# Multiplexed RNA-FISH-guided Laser Capture Microdissection RNA Sequencing Improves Breast Cancer Molecular Subtyping, Prognostic Classification, and Predicts Response to Antibody Drug Conjugates

**DOI:** 10.1101/2023.12.05.23299341

**Authors:** Evan D. Paul, Barbora Huraiová, Natália Valková, Natalia Birknerova, Daniela Gábrišová, Sona Gubova, Helena Ignačáková, Tomáš Ondris, Silvia Bendíková, Jarmila Bíla, Katarína Buranovská, Diana Drobná, Zuzana Krchnakova, Maryna Kryvokhyzha, Daniel Lovíšek, Viktoriia Mamoilyk, Veronika Mančíková, Nina Vojtaššáková, Michaela Ristová, Iñaki Comino-Méndez, Igor Andrašina, Pavel Morozov, Thomas Tuschl, Fresia Pareja, Pavol Čekan

**Author notes:** Corresponding author Pavol Čekan; Fresia Pareja.

## Abstract

On a retrospective cohort of 1,082 FFPE breast tumors, we demonstrated the analytical validity of a test using multiplexed RNA-FISH-guided laser capture microdissection (LCM) coupled with RNA-sequencing (mFISHseq), which showed 93% accuracy compared to immunohistochemistry. The combination of these technologies makes strides in i) precisely assessing tumor heterogeneity, ii) obtaining pure tumor samples using LCM to ensure accurate biomarker expression and multigene testing, and iii) providing thorough and granular data from whole transcriptome profiling. We also constructed a 293-gene intrinsic subtype classifier that performed equivalent to the research based PAM50 and AIMS classifiers. By combining three molecular classifiers for consensus subtyping, mFISHseq alleviated single sample discordance, provided near perfect concordance with other classifiers (κ > 0.85), and reclassified 30% of samples into different subtypes with prognostic implications. We also use a consensus approach to combine information from 4 multigene prognostic classifiers and clinical risk to characterize high, low, and ultra-low risk patients that relapse early (< 5 years), late (> 10 years), and rarely, respectively. Lastly, to identify potential patient subpopulations that may be responsive to treatments like antibody drug-conjugates (ADC), we curated a list of 92 genes and 110 gene signatures to interrogate their association with molecular subtype and overall survival. Many genes and gene signatures related to ADC processing (e.g., antigen/payload targets, endocytosis, and lysosome activity) were independent predictors of overall survival in multivariate Cox regression models, thus highlighting potential ADC treatment-responsive subgroups. To test this hypothesis, we constructed a unique 19-feature classifier using multivariate logistic regression with elastic net that predicted response to trastuzumab emtansine (T-DM1; AUC = 0.96) better than either *ERBB2* mRNA or Her2 IHC alone in the T-DM1 arm of the I-SPY2 trial. This test was deployed in a research-use only format on 26 patients and revealed clinical insights into patient selection for novel therapies like ADCs and immunotherapies and de-escalation of adjuvant chemotherapy.

## Introduction

Breast cancer (BCa) is a heterogeneous disease with distinct biology leading to differences in response to various treatment modalities and clinical outcomes ^1^. The discovery of molecularly distinct subgroups of BCa, i.e., luminal A, luminal B, HER2-overexpressing, basal-like, and normal-like, based on gene expression profiles using microarrays ^2–4^, has fundamentally changed our understanding of BCa biology and paved the way for a union between genomic and clinical classification of BCa subtypes. These intrinsic molecular subgroups markedly differ in terms of prognosis, response to therapy, and clinical outcomes. Within this new framework, various commercially available multigene assays have emerged that can provide important diagnostic, predictive, and prognostic insights to inform them about appropriate treatments ^5,6^.

Assignment of an individual tumor to any subtype (or prognostic risk group), however, shows only moderate reproducibility depending on the array platform used, the tumor composition, gene signature list, and gene expression thresholds ^7–10^. Most multigene tests only provide prognostic and predictive information for ER+/HER2– BCa patients, but not for the more aggressive HER2+ and TNBC subtypes, thus are limited to providing information about who may or may not benefit from chemotherapy in addition to endocrine therapy. ^11^. Current diagnostic tools are also limited in selecting patients that are likely to respond to novel therapeutics such as antibody-drug conjugates (ADCs). The pivotal trials for the ADCs, trasztuzumab deruxtecan (Enhertu) and sacituzumab govitecan (Trodelvy), revealed efficacy in patients that even expressed low to negative levels of their antigen targets when assessed using immunohistochemistry (IHC). This has spurred interest in new tools that can quantify biomarker expression at broad dynamic range and can better stratify individual with low or no expression of an ADC target (e.g., Her2 low). Given their complex mechanisms of action that include not only the antigen target, but also internal processing of the ADC and targeting of the cytotoxic payload, effective patient selection may require assessing multiple biomarkers that are tailored towards the ADC construct.

Multigene assays are largely based on bulk processing or crude macrodissection to enrich for tumor content, thus losing the spatial context, which results in limited info about the tumor microenvironment and also may introduce erroneous gene expression from non-tumor elements present in the bulk-processed specimen ^12^. Indeed, tumor heterogeneity, whether it is histological, biomarker, or genetic, occurs both spatially and temporally and may contribute to diagnostic inconsistencies and inappropriate treatment ^13^. Various tools aiming to combine gene expression data with spatial information have recently been developed ^14^. These emerging spatial biology tools show immense potential in understanding complex BCa biology. Their cost, throughput, evolving chemistry and platforms, and suboptimal trade-off between plexity and resolution pose challenges for their utilization in clinical diagnostics and therapeutic decision making. Thus, laser capture microdissection remains a powerful tool that can ensure tumor purity and can be scaled for clinical diagnostics.

Here, we sought to retrospectively validate the utility of mFISHseq, a diagnostic tool that integrates multiplexed fluorescent *in situ* hybridization (FISH) to assess tumor heterogeneity and guide laser capture microdissection and RNA sequencing. We demonstrate three key features of this approach, including 1) its analytical validity by assessing concordance to gold-standard immunohistochemistry results; 2) a consensus subtyping and prognostic risk approach that mitigates the limitations of individual multigene tools; and 3) leveraging transcriptome profiling of individual gene and gene signature expression to provide clinical insights into prognosis and response to treatments such as ADCs.

## Results

### Study design and cohorts

We conducted the retrospective clinical validation of mFISHseq, a diagnostic tool that uses a multiplexed RNA-FISH panel consisting of estrogen (*ESR1*), progesterone (*PGR*), and HER2 (*ERBB2*) receptors and Ki67 (*MKI67*) to characterize regions of interest that are subsequently captured by laser capture microdissection (mFISHseq). This facilitates the collection of spatially resolved, tumor-enriched samples from a single specimen that can be used for downstream total RNA sequencing (**Figure 1a**), providing a powerful tool to interrogate tumor heterogeneity and biology. We conducted this analysis on a cohort of 1,082 FFPE BCa samples with detailed clinicopathological information (**Figure 1b, Extended Table 1**). We first assessed its analytical validity by comparing results with the known biobank IHC status. Then we generated a consensus molecular subtyping approach by using a research based PAM50, Absolute Intrinsic Molecular Subtyping (AIMS), and our own subtyping scheme and investigated its performance in stratifying samples according to prognostic risk. Finally, we demonstrated its clinical potential by assessing the expression of genes and gene signatures with diagnostic, prognostic, and predictive value.

**Figure 1.**
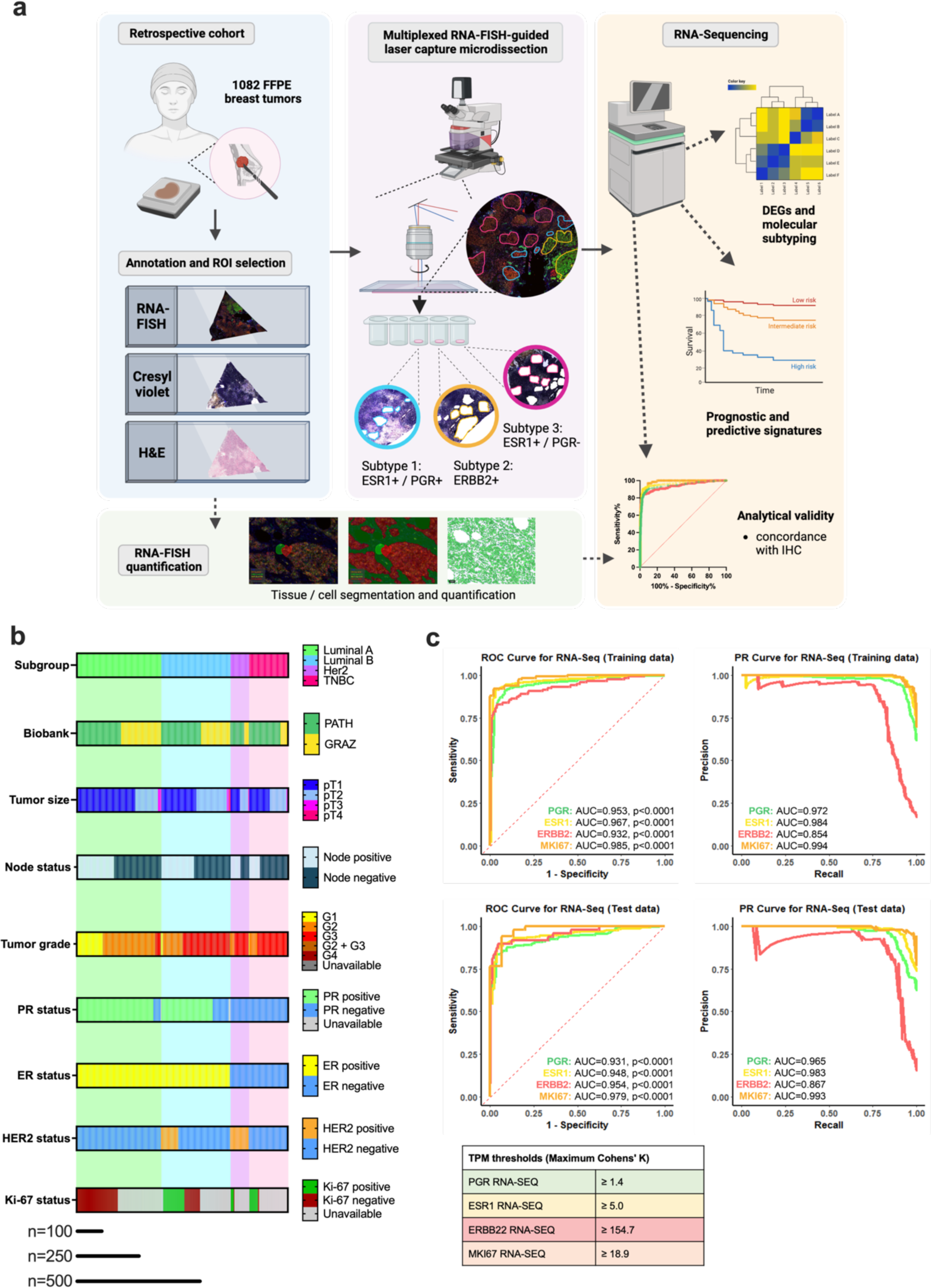
Study design, workflow, and analytical validity. (**a**) The retrospective validation cohort consisted of 1,082 formalin-fixed paraffin-embedded (FFPE) breast cancer samples, which underwent multiplexed RNA-FISH-guided laser capture microdissection (LCM) coupled with RNA-sequencing. Annotation of the tumor area on an H&E section and the biomarker expression derived from multiplexed RNA-FISH were used to select regions of interest (ROIs) for LCM from cresyl violet sections. These tumor-enriched samples were then sequenced to characterize gene expression signatures to provide diagnostic, prognostic, and predictive inferences from the cohort clinical data (Created with BioRender.com) (**b**) Clinicopathologic features of the retrospective cohort according to intrinsic molecular subtype. (**c**) Analytical validity of mFISHseq compared to immunohistochemical data as assessed by receiver operating characteristic (ROC) and precision-recall (PR) curves in 70:30 training and test datasets. (**d**) Individual biomarker thresholds defined in the training set and applied in the test set.

### Concordance with biobank immunohistochemistry status

To assess the analytical validity of the mFISHseq assay, we compared the results from RNA-FISH and -SEQ to the ER, PR, HER2, and Ki67 IHC results reported by the biobanks. Both RNA-FISH and RNA-SEQ showed similar expression patterns compared to IHC for PR, ER, HER2, and Ki67 (**Supplementary Figure 1**), especially at high expression levels, and recapitulated the expression patterns when divided into clinical molecular subtypes (**Supplementary Figure 2**). To determine appropriate threshold values for mFISHseq, we split our dataset into training and test cohorts (70:30) and constructed receiver operating characteristic (ROC) and precision-recall (PR) curves in comparison to the known IHC results, we observed that RNA-SEQ and IHC had excellent concordance with ROC and PR curves with all biomarkers having ROC and PR AUCs > 0.90, with the exception of the PR curves for *ERBB2*, which showed PR AUCs > 0.85 on both training and test data (**Figure 1c, Supplementary Table 1**). While RNA-FISH was primarily used to guide LCM, we used a digital pathology and machine learning pipeline (**Extended Figure 1**) to quantify fluorescent intensity within tumor cells and found very good concordance with the known biobank IHC results (ROC and PR curves ≥ 0.80 for all biomarkers; **Supplementary Figure 3a-d**), suggesting both orthogonal methodologies have diagnostic utility. Overall, these results are consistent with prior reports that have assessed the performance of RNA-FISH/-SEQ when compared with IHC ^15,16^ and thus support the analytical validity of the mFISHseq methodology in assessing classical IHC breast cancer biomarkers.

### Cross-validation of RNA-FISH and RNA-SEQ

Using two independent techniques to assess biomarker expression allows cross-validation of the results from each technique, consequently mitigating false negatives/positives that arise by using a single method. Both techniques show mild to very strong correlations for the detection of each biomarker, with *ESR1* (r=.75) having the strongest correlation followed by *PGR* (r=.66), *MKI67* (r=.61), and *ERBB2* (r=.41) (**Supplementary Figure 4a-d**). The lower correlation for *ERBB2* is driven by the large number of *ERBB2* negative samples (4:1 negative to positive ratio) since the correlation between RNA-FISH and RNA-SEQ was expectedly poor for negative samples (r=.17) whereas the positive samples were very strongly correlated (r=.69). According to a Bland Altman analysis on normalized Z-scores, both techniques showed good agreement and low bias according to 95% agreement intervals with most bias occurring when targets were expressed at low levels (**Supplementary Figure 4e-h**).

### Benchmarking of mFISHseq molecular subtypes with IHC surrogates, AIMS, and PAM50

To identify gene signatures that can effectively classify breast cancer specimens into the four intrinsic molecular subtypes, we conducted differential gene expression analyses using DeSeq2 and the Wilcoxon rank-sum test ^17^, a nonparametric approach that better controls the false discovery rate (FDR) when analyzing large sample sizes. With these differentially expressed genes (**Extended Table 2**), we generated a list of 293 genes (see **Supplementary Dataset 1** and **Supplementary Methods**) and applied semi-supervised consensus clustering to determine how these samples clustered according to the original biobank subtype designation (**Figure 2a**). The top 5 upregulated and downregulated genes (log2 fold change >2, and Wilcoxon FDR < 0.05) for each subtype comparison can broadly be segregated into genes involved in luminal pathways (*ESR1*, *GATA3, CA12, THSD4*), genes coamplified with *ERBB2* (i.e., Her2 amplicon - *ERBB2*, *GRB7*, *PGAP3*, and *MIEN1*), basal epithelial markers (*KRT16*, *KRT81*, and *SOX10*), and proliferation markers (S100 genes)(**Supplementary Figure 5a**). As a benchmark for RNA-based gene expression signatures, we also called PAM50 subtypes and Absolute Intrinsic Molecular Subtyping (AIMS) using Genefu ^18^ as well as TNBCtype ^19^ to classify Lehmann’s six and four TNBC subtypes ^20,21^.

**Figure 2.**
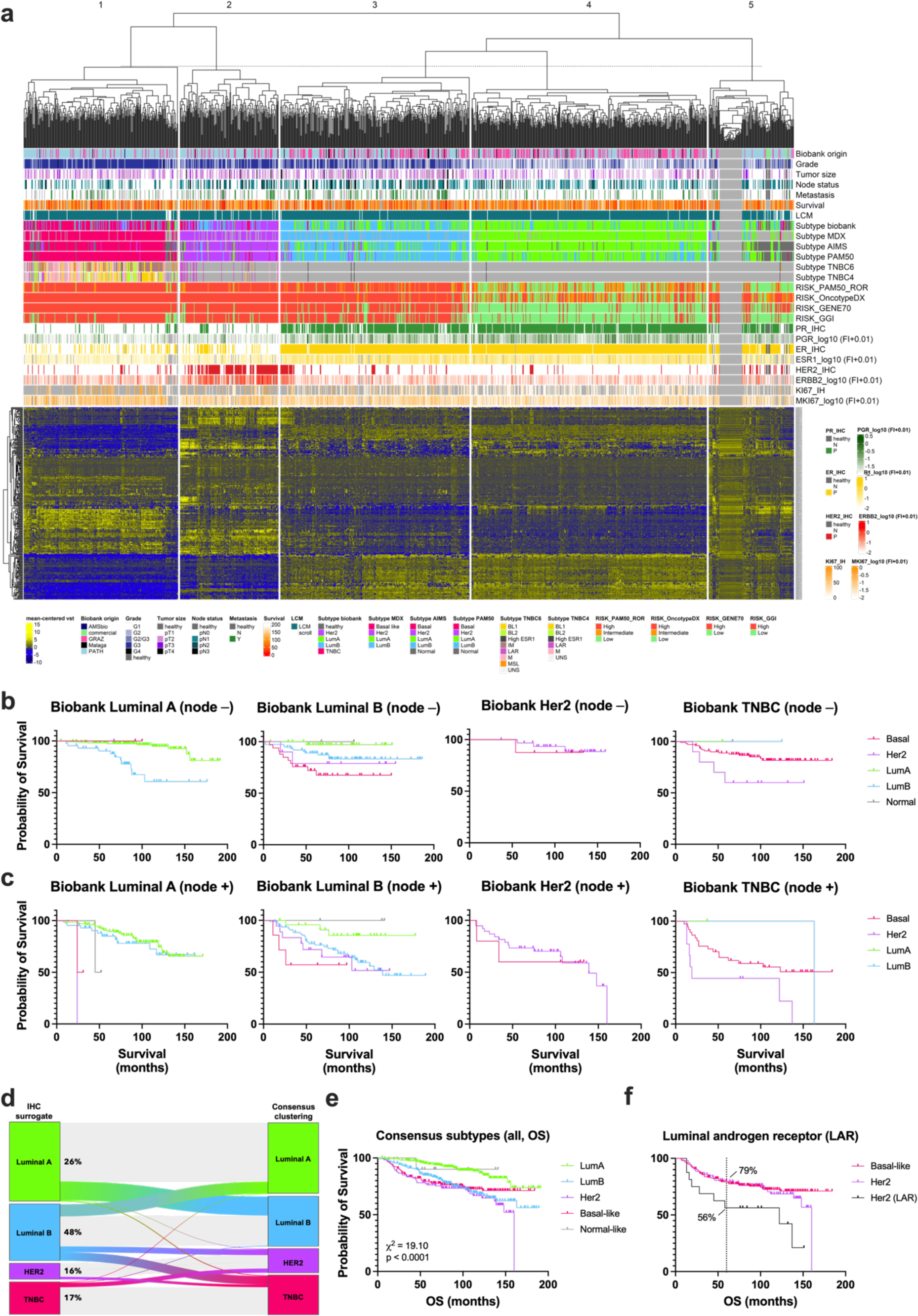
Consensus subtyping yields intrinsic molecular subtypes associated with survival. (**a**) Gene expression heatmap depicting the 293 differentially expressed genes in 1013 breast breast cancer samples used for molecular subtyping. The metadata shown in phenobars above the gene expression heatmap includes (from top to bottom) biobank origin, clinical parameters, LCM or no LCM, subtyping approaches, prognostic signatures, and IHC and RNA-FISH results. (**b**,**c**) Overall survival according to consensus subtyping with respect to their clinical subtype according to the biobank IHC results and nodal status. (**d**) The Sankey diagram shows the clinical subtypes according to biobank IHC classification and the proportion of samples reclassified by mFISHseq consensus clustering. (**e**) Overall survival of consensus molecular subtypes. (**f**) Overall survival of the Her2 consensus molecular subtype samples classified as the luminal androgen receptor (LAR) TNBC subtype in relation. To other non-LAR Her2 and basal-like consensus subtype samples. The vertical dashed line and annotated percentages denote probability of overall survival at 60 months.

Each multigene subtyping method classified fewer luminal A samples and more Her2 and basal-like samples relative to the IHC surrogate subtypes. When comparing multigene classifiers, AIMS had the most luminal A, Her2, and normal samples and least basal-like samples. The mFISHseq and PAM50 subtyping approaches had the most luminal B and basal-like samples, and PAM50 classified the fewest luminal A samples (**Supplementary Figure 5b**). While all subtyping methods showed similar overall survival curves (**Supplementary Figure 5c**), there was only moderate concordance between each multigene classifier and the IHC surrogate subtype at the single sample level (Cohen’s kappa (κ): mFISHseq=0.567, PAM50=0.539, AIMS=0.519; **Supplementary Figure 5c**), with the luminal B subtype showing only fair agreement (κ range = 0.36-0.384), while the basal-like/TNBC subtypes had substantial agreement (κ range = 0.638-0.704). In contrast to prior reports ^7,8^, we found substantial concordance between multigene approaches with MDX293 vs PAM50 showing the highest concordance (κ=0.748). For all comparisons between multigene classifiers, the luminal B subtype again showed the lowest agreement (κ range = 0.631-0.683) followed by luminal A (κ range = 0.668-0.722), Her2 (κ range = 0.773-0. 837), and basal-like with the best agreement (κ range = 0.891-0.968).

For samples that showed high levels of discordance between multigene subtyping methods and IHC surrogate subtypes (i.e., at least 2-3 classifiers providing discordant results), we observed clinically relevant differences in survival (**Supplementary Figure 5e**). IHC surrogate luminal A samples that had discordant results showed poorer survival than samples that were classified as luminal A by all multigene classifiers (**Supplementary Figure 5e**; top panel). In contrast, IHC surrogate luminal B samples that had discordant results by two classifiers had longer survival than samples that were classified as luminal B by all multigene classifiers (**Supplementary Figure 5e**; middle-top panel). While discordant Her2 samples showed comparable survival to concordant Her2 samples (i.e., those classified as Her2 by all approaches; **Supplementary Figure 5e**; middle-bottom panel), discordant TNBC samples interestingly showed even poorer survival than concordant TNBC samples (**Supplementary Figure 5e**; bottom panel). In general, discordant samples, relative to concordant samples, showed evidence of instability as demonstrated by lower correlations to PAM50 centroids. The correlations to the PAM50 centroid were higher for samples where all four classifiers (IHC surrogate, mFISHSeq, AIMS, and PAM50) agreed on the classification and progressively decreased for discordant samples where one, two, or all three classifiers disagreed on the subtype classification (**Supplementary Figure 5f**).

### Consensus subtyping improves concordance at the single sample level and mitigates misclassification

To further improve agreement at the sample level, we constructed a consensus intrinsic subtype by using a simple voting scheme for the three subtyping approaches (mFISHseq, PAM50, and AIMS) and found there were considerable differences between IHC surrogate and gene-expression based consensus subtypes that had prognostic implications. For IHC surrogate luminal A samples, 45% (193/432) showed discordance in one or more alternative classification schemes, resulting in 24% (102/432) of samples being reclassified as luminal B, which also showed poorer overall survival for node negative, but not node positive samples, relative to samples unanimously classified as luminal A by all subtyping methods (**Figure 2b,c** left panel). A small number of IHC luminal A samples (10/432 or 2%) were reclassified as either Her2 (n=2), basal-like (n=4), or normal-like (n=4); however, the numbers were too small to make conclusions regarding overall survival, The IHC surrogate luminal B subtypes showed high discordance with 62% (194/313) disagreeing with one or more classifiers. Around 15% (46/313) of samples were reclassified into the basal-like subtype and 21% (65/313) into the luminal A subtype by consensus subtyping and these patients had either poorer or more favorable survival, respectively, compared to samples unanimously classified as luminal B (**Figure 2b,c** middle-left panels). The 10% (32/313) of IHC luminal B samples reclassified as Her2 by consensus subtyping had similar survival to consensus luminal B samples (**Figure 2b,c** middle-left panels). We observed the least discordance in the Her2 IHC surrogate samples where 27% (20/74) displayed disagreement among classifiers, resulting in reclassification of 19% (14/74) of samples as the basal-like subtype, which did not differ in overall survival from consensus Her2 samples (**Figure 2b,c** middle-right panels). Disagreement among classifiers occurred in 29% (53/181) of TNBC IHC surrogate samples, resulting in reclassification of 4% (7/181) of samples into luminal A and B and normal-like subtypes as well as 13% (23/181) of samples into the Her2 subtype, and these reclassified samples showed more favorable and poorer survival, respectively, when compared to consensus basal-like samples (**Figure 2b,c** right panels).

We next explored the samples that were classified uniquely by a single classifier to ascertain the degree of discordance and clinical implications at the single sample level. Each approach uniquely classified a portion of samples as luminal A that no other classifier agreed upon, and these samples uniquely classified by IHC surrogate (n=67), mFISHseq (n=15), AIMS (n=25), and PAM50 (n=2) showed poorer overall survival relative to the 239 samples classified as luminal A by all tools (LumA All; **Supplementary Figure 6a**). While LumA All samples were enriched in low/intermediate prognostic risk scores (assessed by research-based versions of OncotypeDX, GENE70, ROR-S, and clinical risk), the uniquely classified samples had high genomic and clinical risk (**Supplementary Figure 6b**), suggesting that each subtyping method is susceptible to misclassifying a portion of samples. These misclassified samples, however, were ‘rescued’ by taking the consensus of the three multigene classifiers with 83% (n=91/109) being reclassified as luminal B (**Supplementary Figure 6b**, bottom right panel). In contrast, samples that were uniquely classified as luminal B showed more favorable overall survival relative to samples classified as luminal B by all tools (LumB All), except for the IHC surrogate approach, which showed similar survival (**Supplementary Figure 6c**). The samples uniquely classified as luminal B by mFISHseq (n=29) and PAM50 (n=42) contained more samples with intermediate/low risk scores, relative to LumB All samples (**Supplementary Figure 6d**), and 97% (n=69/71) of these were reclassified as luminal A by consensus subtyping (**Supplementary Figure 6d**, bottom right panel). Notably, AIMS did not uniquely classify any luminal B samples. The samples classified as luminal B only by the IHC surrogate approach were reclassified by consensus subtyping into luminal A (n=29)/normal-like (n=6) subtypes and Her2 (n=24)/basal-like (n=46; **Supplementary Figure 6d**, bottom right panel), which showed either more favorable or poorer overall survival, respectively, relative to LumB All samples (**Supplementary Figure 6c**, inset).

The samples classified as Her2 by only a single classifier showed disparate findings depending on the classifier (**Supplementary Figure 6e,f**). IHC surrogate Her2 samples (n=14) had equivalent overall survival relative to Her2 All samples (n=67), high genomic and clinical risk, and were unanimously reclassified as basal-like by consensus subtyping. AIMS Her2 samples (n=24) had poorer overall survival relative to Her2 All samples, high genomic and clinical risk, and nearly all samples were reclassified as either basal-like (n=15/24) or luminal B (n=8/24). The mFISHseq (n=9) and PAM50 (n=14) Her2 samples generally had more favorable survival and a subset of these samples (n=9) with low genomic (GENE70) and/or clinical risk were reclassified by consensus subtyping into predominantly luminal A (n=7/9) and had no events.

TNBC and the similar basal-like samples showed the least amount of uniquely identified samples, with only 28 TNBC samples uniquely identified by IHC surrogate subtyping, and six basal samples uniquely classified by mFISHseq (n=2) and PAM50 (n=4), while AIMS did not independently classify any basal-like samples. While IHC surrogate only TNBC samples showed poorer overall survival compared to the 128 samples classified as TNBC/basal-like by all methods (Basal-like All), the samples classified as basal-like by the multigene classifiers had better prognosis (**Supplementary Figure 6g**). Interestingly, IHC surrogate only TNBC samples could be stratified into 13 samples that had either low GENE70 genomic or clinical risk and showed favorable survival, while the other 15 samples had high risk and poor survival (**Supplementary Figure 6g**, inset, **6f**). Consensus subtyping was also capable of stratifying the samples into clinically meaningful subgroups with 10/22 samples reclassified as Her2 or basal-like dying, while only 1/5 samples reclassified as luminal A or B died (**Supplementary Figure 6f**). We also note that AIMS independently classified 34 samples as normal-like, while PAM50 independently classified only 1 sample (12 samples were classified as normal-like by both classifiers), and these were reclassified by consensus subtyping to basal-like (n=18), luminal A (n=14), and Her2 (n=3), thus providing more clinically relevant subtyping (**Supplementary Figure 7a,b**).

Consensus subtyping was able to reclassify 214 samples uniquely classified by the IHC surrogate methodology into intrinsic subtypes that better fit the survival data. Similarly, the samples that were uniquely classified by mFISHseq (n=55), AIMS (n=83), and PAM50 (n=63) were all reclassified by a consensus of the other two multigene classifiers, which yielded more reproducible assignment at the single sample level to the appropriate intrinsic subtype that matched the overall survival. Thus, consensus subtyping effectively mitigates one of the impediments to adopting molecular subtyping as a clinically useful tool, namely the irreproducibility of classifying single samples with a single approach.

Overall, 30% (305/1013) of samples were reclassified from their IHC surrogate subtype into a different consensus subtype with luminal B samples being reclassified the most (48%, 149/313) followed by luminal A (26%, 112/432), TNBC (17%, 30/181), and Her2 samples (16%, 14/87; **Figure 2d**). Kaplan-Maier analysis showed that consensus subtypes have prognostic utility with 5-year and 10-year overall survival rates of 94.4%, 84.3%, 76.3%, and 79.4% and 86.7%, 68.3%, 66.8%, and 72.7% for luminal A, luminal B, Her2, and basal-like subtypes, respectively (**Figure 2e**). Interestingly, a portion of IHC TNBC samples that were reclassified as Her2 clustered as the luminal androgen receptor subtype according to TNBCtype ^19^ and these samples had poorer survival than other Her2 and TNBC/basal-like samples with 5-year overall survival rates of only 56.3% compared to 79.3% for consensus Her2 and TNBC/basal-like samples (**Figure 2f**). Given the discordance in molecular subtyping approaches we and others have observed, the consensus molecular subtyping scheme also alleviated misclassification at the single sample level, which resulted in better stratification of individuals into poor and favorable prognosis than any one subtyping scheme on its own. This improved concordance between the consensus subtypes and other multigene classifiers to near perfect agreement, even for more challenging luminal samples (**Supplementary Figure 5d**)

### Consensus prognostic risk categories alleviates discordance at the single-patient level and identifies patients with ultra-low risk

We also investigated the performance and concordance of several multigene prognostic risk assays since these tests can provide information on the effectiveness of adjuvant chemotherapy in select ER+/Her2− patients with 0-3 positive lymph nodes. We compared clinical risk, as assessed using criteria in the MINDACT trial, to research-based versions of OncotypeDX, the PAM50 Risk of Recurrence by Sample (ROR-S), GENE70 (i.e., MammaPrint), and the Genomic Grade Index (GGI). This analysis was restricted to 567 patients that were ER+/Her2− by IHC and either node negative (pN0) or node positive (1-3 positive nodes, pN1), encompassing the indicated population of patients eligible for the commercial tests. Relative to clinical risk, all multigene assays classified fewer patients as high risk with OncotypeDX and GGI having the highest and lowest proportion of high-risk patients, respectively. GENE70 and GGI, which use a 2-category (high and low) risk assessment scale, classified more patients as low risk, relative to clinical risk, OncotypeDX, and ROR-S (**Figure 3a**).

**Figure 3:**
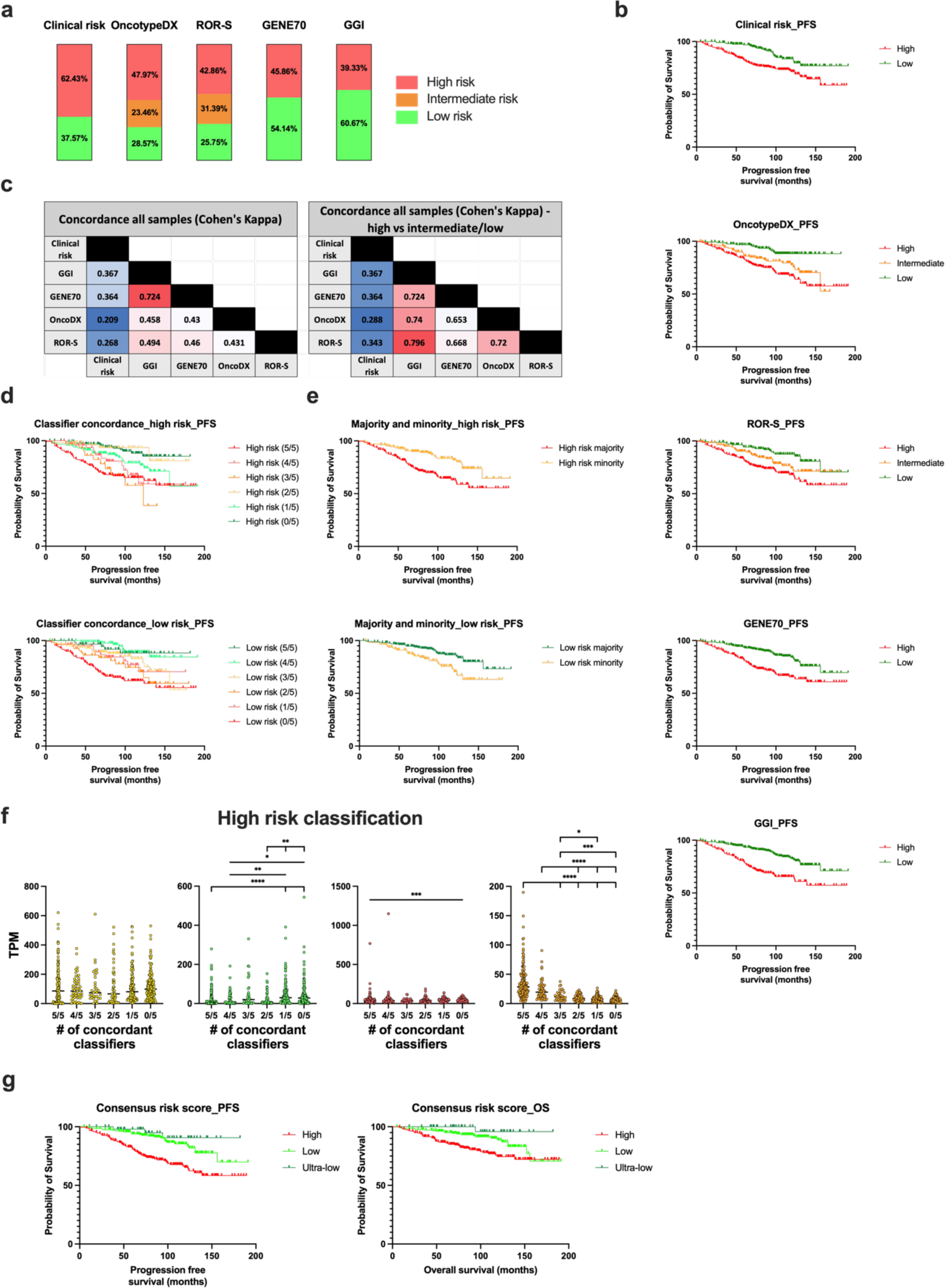
**Consensus prognostic risk categories show clinically relevant differences in survival** (a) Proportion of risk categories (high, intermediate, and low) among each of the five prognostic classifiers, including Clinical risk, OncotypeDX, Risk of Recurrence by Subtype (ROR-S), GENE70, and Genomic Grade Index (GGI) on 567 ER+/HER2− with 0-3 positive lymph nodes. (b) Progression free survival (PFS) of each of the five prognostic classifiers. (**c**) Concordance of each classifier when comparing all risk categories (high, intermediate, and low; left table) or after consolidating into two categories (high and intermediate/low combined; right table). (**d**) Concordance of each classifier for high and low risk samples as illustrated by the number of concordant classifiers and (**e**) after consolidating into majority (agreement in ≥3 prognostic classifiers) and minority (agreement in 1-2 prognostic classifiers) categories. (**f**) Distribution of mRNA expression for estrogen receptor (*ESR1*, yellow, left panel), progesterone receptor (*PGR*, green, left/middle panel), HER2 receptor (*ERBB2*, red, right/middle panel), and Ki67 marker of proliferation (*MKI67*, orange, right panel) in patients that were classified as high risk by a particular number of concordant classifiers (i.e., 1-5 concordant classifiers). (**g**) Kaplan Meier plots show PFS and overall survival (OS) for each consensus prognostic risk category (high, low, and ultra-low).

All risk classifiers had comparable prognostic utility with low- and high-risk patients showing favorable and poor progression-free survival (PFS), respectively (**Figure 3b**). Agreement between multigene classifiers and clinical risk was only fair (κ: GGI=0.367, GENE70=0.364, ROR-S=0.343, OncotypeDX=0.288), while agreement among multigene classifiers was moderate to substantial (κ range: 0.431 - 0.724), with GENE70 and GGI showing the highest agreement and GENE70 and OncotypeDX having the lowest (**Figure 3c**). Since OncotypeDX and ROR-S use a 3-category (high, intermediate, low) risk scale and patients classified as intermediate by these two classifiers showed only slight agreement (κ=0.112) we dichotomized the 3-categories into 2 categories (high vs intermediate/low) improved concordance, especially among multigene classifiers (κ range: 0.653 - 0.796; **Figure 3c**). Only 39.2% of patients (n=222/567) were unanimously classified by all five approaches as either high (n=162) or low (n=60) risk leaving 61.8% of patients (n=345) with a discordant result in at least one classifier. Within these discordant patients, another 27.5% of patients (n=156/567) were categorized as either high (n=52) or low (n=104) by four classifiers and 23.5% of patients (n=133/567) were categorized as either high (n=33) or low (n=100) by three classifiers. Thus, the five prognostic classifiers reached a majority consensus (i.e., at least 3 classifiers in agreement) to stratify 43.6% (n=247/567) and 47.1% (n=267/567) of patients into high and low risk, respectively.

Notably, patients differed markedly in terms of outcome depending on the number of concordant classifiers for a particular risk category (**Figure 3d**). Patients with at least four classifiers in agreement for either high or low risk showed the poorest and best probability of PFS at 10 years (59.2 % for 4/5 classifiers - 65.1% for 5/5 classifiers in agreement for high-risk vs 89.0% for 4/5 classifiers - 88.2% for 5/5 classifiers in agreement for low-risk; **Figure 3d**). When separating patients based on whether a majority (3 or more classifiers in agreement) or minority (1 or 2 classifiers in agreement) of the five classifiers predicted the same risk category, we observed that the minority group displayed PFS that was intermediate compared to the majority as well as patients classified unanimously as high or low risk (**Figure 3e**). Patients classified as high risk by the majority of classifiers had lower expression of PGR and higher MKI67 mRNA relative to patients classified as high risk by the minority of classifiers, while there was no difference in ESR1 expression (**Figure 3f**). Patients unanimously classified as high risk also had higher ERBB2 expression (e.g., ERBB2 low) relative to patients that were not classified as high risk by any classifier (**Figure 3f**). Each classifier independently categorized a proportion of patients into high (n=120), intermediate (n=186), and low (n=65) risk while no other classifier agreed with these unique classifications (**Supplementary Figure 8a**). Clinical risk had the most unique classifications (n= 118) followed by ROR-S (n=104), OncotypeDX (n=103), GGI (n=25), and GENE70 (n=21). Uniquely classified intermediate and high-risk patients had better progression free survival relative to patients classified as intermediate risk by both OncotypeDX and ROR-S and high risk by all classifiers, respectively (**Supplementary Figure 8b**). The better survival for the multigene assays was more evident when each classifier was combined because of the low patients per classifier (**Supplementary Figure 8c**). Patients independently categorized as low risk by GGI or GENE70 had poorer survival and those uniquely classified as low risk by Clinical risk were similar relative to patients classified as low risk by all classifiers (**Supplementary Figure 8b,c**).

Given the discordance observed at the single-patient level for risk categorization, we constructed consensus prognostic risk categories by combining the results for all five classifiers into 3-risk categories: high risk (If ≥3 prognostic classifiers agree on intermediate/high risk), low risk (If ≥3 prognostic classifiers agree on low risk), and ultra-low risk (If all 5 prognostic classifiers agree on low risk and the patient is node negative (pN0) and PR+) as outlined in the decision tree in **Supplementary Figure 8d**. Consensus high risk patients (n=300) showed poor outcomes (PFS and OS) with 49 relapses (distant and local) and 36 deaths within 5-years and another 24 relapses and 17 deaths from 5-10 years. Consensus low risk patients (n=214) had better outcomes with 13 relapses and 10 deaths within 5-years and another 10 relapses and 7 deaths from 5-10 years. Consensus ultra-low risk patients (n=53) showed the best outcomes with 1 relapse and 0 deaths within 5-years and another 2 relapses and 1 death from 5-10 years (**Figure 3g**). When stratifying the consensus outcome groups based on treatment (**Supplementary Figure 8e**), we observed that high risk patients benefited most from chemoendocrine therapy, relative to endocrine or chemotherapy alone, while low/ultra-low patients did not benefit from chemoendocrine therapy, highlighted the clinical utility of our consensus prognostic risk categories in de-escalating overtreatment in low/ultra-low risk patients.

### Interrogation of genes and gene signatures to identify prognostic subgroups that may predict therapeutic response

To investigate potential markers of prognosis and response to treatment, we curated a list of 92 genes and 110 gene signatures and assessed their expression in our cohort (n=1013) in relation to BCa IHC surrogate subtype and overall survival (**Supplementary Dataset 2**). Supervised clustering of these signatures largely segregated samples into their respective molecular subtypes, although Her2 enriched samples formed a cluster with luminal B samples that contained amplified Her2 (assessed by ICH/FISH) or elevated *ERBB2* expression (**Figure 4a**).

**Figure 4.**
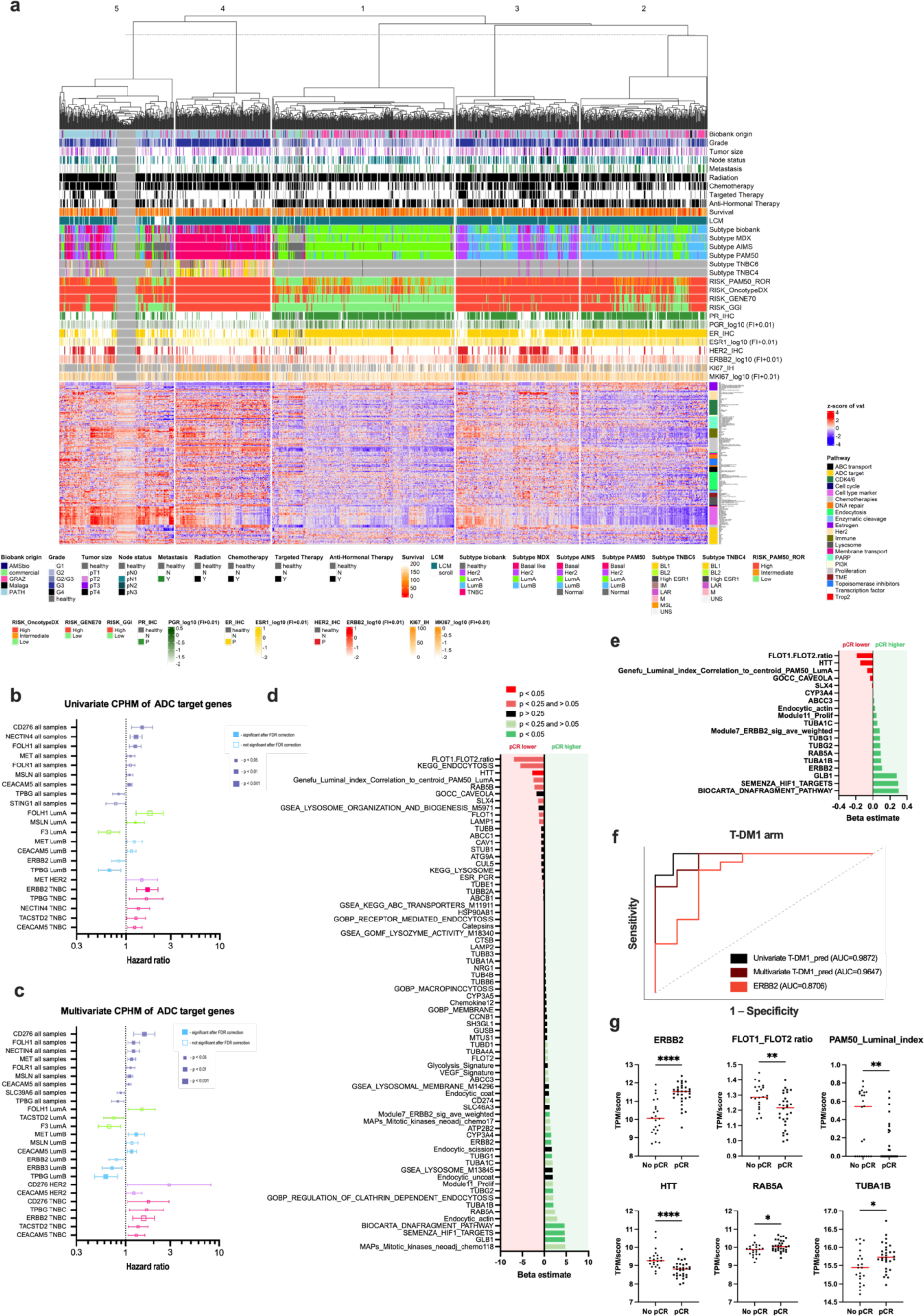
Gene signatures associated with survival and prediction of treatment response. (**a**) Gene expression heatmap illustrating the expression of 92 genes and 110 gene signatures in respect to molecular subtype and clinical parameters. Univariate (**b**) and multivariate (**c**) Cox proportional hazards models (CPHM) on 20 ADC targets and their association with overall survival in all samples and stratified by subtype. (**d**) Univariate logistic regression analysis of 70 genes/gene signatures and their association with pathologic complete response (pCR) in the T-DM1 arm of the I-SPY2 trial. (**e**) The 19 genes/gene signatures selected in the multivariate logistic regression with elastic net modeling on the training dataset. Green bars denote signatures associated with pCR; red bars indicate signatures associated with no pCR. (**f**) Performance of two T-DM1_pred classifiers in the test set relative to ERBB2 alone. Univariate T-DM1_pred is a single score derived from all 19 features, while multivariate T-DM1_pred includes all 19 features in a multivariate regression model. AUC = area under the curve. (**e**) Scatter plots showing the distribution of selected genes/gene signature scores of the T-DM1_pred classifier in patient samples according to pCR.

A univariate Cox analysis on all samples, irrespective of IHC subtype, revealed the signatures associated with poor survival were the hypoxia/angiogenesis (Metabolic_sig_Trop2_TNBC_Survival, VEGF_Signature, Glycolysis_Signature, Hypoxia_Angiogenesis_Inflammatory_MDX), prognostic scores (PAM50 ROR-S, GENE70, OncotypeDX, and GGI), proliferation markers (*AURKA*, *CCNB1, CCNE1*, *FOXM1*, *MKI67*, *TK1*, PMID_17493263_proliferation, Module11_Prolif, and Proliferation_MDX), and DNA damage repair (*RAD51*, *TK1*), while signatures associated with favorable outcome included luminal (*PGR*, TAMR13_scores, MOTERA, Genefu_Luminal_index_Correlation_to_centroid_PAM50_LumA, and GSEA_HALLMARK_ESTROGEN_RESPONSE_EARLY) and immune (Mast_cells; **Extended Figure 2, Supplementary Dataset 3**) pathways. The prognostic scores and some proliferation markers were also associated with poor survival in luminal A and B subtypes, but not Her2 or TNBC (**Extended Figure 3, Supplementary Dataset 3**). The hypoxia/angiogenesis signatures predicted poor survival in luminal B and TNBC, albeit in the latter the significance did not remain after FDR correction. Different immune signatures were prognostic for different subtypes. While dendritic cell and neutrophil signatures were associated with poor prognosis in luminal A and luminal B (not FDR corrected), signatures related to dendritic cells, monocytes/macrophages, and T and B cells were associated with a favorable prognosis in TNBC samples (not FDR corrected).

Nearly all the significant signatures in the univariate analysis on all samples remained independent predictors of survival when including both tumor size (pT1 vs pT2-4) and node status (pN0 vs pN1-3) into multivariate Cox models (**Supplementary Dataset 4**), even when adjusting for multiple hypothesis testing. In contrast, analyzing the signatures at the subtype level yielded fewer independent predictors, especially when using FDR correction. We observed no independent predictors in luminal A, only a single predictor for Her2 (BIOCARTA_DNAFRAGMENT_PATHWAY associated with favorable survival), and only two predictors of poor survival for TNBC (Module7_ERBB2_sig_ave_weighted and Pathologic_response_ER.Neg) when FDR correcting for multiple tests. The exception was luminal B where we observed additional independent predictors that were apparently confounded during the univariate analysis, including signatures related to luminal pathways (GSEA_HALLMARK_ESTROGEN_RESPONSE_EARLY, MOTERA_sig_ESR1_mutations_Cancer_Res_2021, ESR_PGR_ave, PMID_17493263_luminal_epithelial, and TAMR13_scores), immune function (Mast_cells), and microtubules (*MTUS1*) associated with favorable survival as well as signatures related to chemotherapy (HS_Red1, MAPs_Mitotic_kinases_neoadj_chemo118), cell cycle (cMET_FAK_CDK2_axis), and Her2 resistance (*MUC4*) associated with poor survival. There were also several ADC targets that were independent predictors of survival that were not significant in the univariate analysis (*CEACAM5*, *MET*, *ERBB3*).

### ADC processing-related genes and gene signatures show subtype-specific expression and association with survival

We further characterized the expression of 20 targets of ADCs that are either approved or undergoing clinical trials and associated their expression with survival. Most ADC targets displayed high inter-individual variability, differential expression between healthy and invasive tumor tissue, and enrichment in a subtype-specific manner (**Extended Figure 4**). Moreover, in both univariate and multivariate analyses (**Figure 4b,c**), many ADC targets were associated with poor survival like *CD276*, *FOLH1*, *NECTIN4*, *MET*, *FOLR1*, *MSLN*, and *CEACAM5* or favorable survival like *SLC39A6* (multivariate only), *TPBG*, and *STING1* (univariate only; **Figure 4b**). The ADC targets were also related to survival in a subtype specific manner and occasionally in opposite ways, which could have important implications for effective patient selection. For example, *TACSTD2*, the gene encoding the TROP2 protein, which is targeted by sacituzumab govitecan (and datopotamab deruxtecan in a phase 3 trial), was associated with favorable survival in luminal A but poor survival in TNBC. Likewise, expression of ERBB2 (the target of trastuzumab deruxtecan, trastuzumab emtansine, and many other ADCs in development), ERBB3 (the target of patritumab deruxtecan), and TPBG predicted favorable survival in luminal B samples but poor survival in TNBC (**Figure 4b,c**).

Because the sensitivity and resistance to ADCs likely encompasses broad cellular targets and pathways involved in ADC processing, we also characterized the expression of genes and gene signatures related to receptor endocytosis, lysosomal function, payload targets (e.g., topoisomerases and microtubules, DNA damage repair), and resistance pathways (multi-drug resistance transporters and metabolizing enzymes). The overarching hypothesis is that combining these signatures may elucidate ADC-responsive patient subgroups that can be exploited for effective patient selection. The expression of these ADC processing related genes/gene signatures displayed marked variability across cases (**Figure 4a**), suggesting that individual variation in the cellular processes involved in the action of ADCs may predict response to these agents. Like the ADC targets, other components of ADC processing showed strong association with survival even in multivariate models (**Supplementary Dataset 3**,**4**). In a multivariate analysis of all samples, signatures related to drug resistance/membrane transport (*ABCC1*, GSEA_KEGG_ABC_TRANSPORTERS_M11911), endocytosis (*ATG9A*, *FLOT2*, Endocytic_uncoat, GSEA_GOBP_MEMBRANE_RAFT_ASSEMBLY, Endocytic_actin, Endocytic_coat), lysosome function (*LAMP1*, GSEA_KEGG_LYSOSOME_M11266, GSEA_LYSOSOME_M13845) and enzymatic cleavage (*CTSB*, Cathepsins), and topoisomerase inhibitors (*PROM1*, *RAD51*) were all independent predictors of poor survival (**Supplementary Dataset 4**). This supports the notion that patients could be stratified into subgroups based on the combination of ADC-relevant genes and gene signatures.

We sought to investigate whether genes and gene signatures involved in ADC function could predict response to ADCs. Given that our retrospective validation cohort did not encompass patients treated with these agents, we conducted a re-analysis of the trastuzumab emtansine (T -DM1) arm of the I-SPY2 trial to test our hypothesis. We divided the dataset into a training and a test cohort (50:50) and conducted a univariate logistic regression on a panel of 70 pre-specified ADC relevant genes and gene signatures. Our analyses found that various gene signatures related to different steps of ADC processing (i.e., antigen, receptor internalization, lysosomal proteolysis, payload target, and drug metabolism) were predictive of pathologic complete response (pCR; **Figure 4d**). A multivariate logistic regression model using elastic net and 10-fold cross validation yielded a final 21-feature classifier (**Figure 4e**) that displayed superior predictive utility to *ERBB2* alone (ROC AUC of 0.94 vs 0.87, **Figure 4f**). The dominant features in the signature predicting T-DM1 efficacy were found to be related to the target antigen (*ERBB2*), internalization of the ERBB2 receptor (*FLOT1/FLOT2* ratio, RAB5A), lysosome function (*GLB1*, *HTT*), microtubule targets of the maytansine payload (*TUBA1B*, *TUBA1C*, *TUBG1*, *TUBG2*), and markers of resistance (e.g., the multidrug resistance transporter, *ABCC3*; **Figure 4g**). Altogether these data support the notion that genes and gene signatures that encompass ADC processing and activity can be used to predict response with higher accuracy than the antigen target alone.

### Unraveling the tumor biology of Her2-low

The recent approval of the antibody drug conjugate trastuzumab deruxtecan (Enhertu) for treatment of individuals with low levels of Her2 in the advanced/metastatic setting has spurred lively debate on whether Her2 constitutes a unique biological entity^22^. Rather than assessing Her2 low in the context of IHC results, which are broadly acknowledged to poorly separate low levels of Her2 from the absence of Her2 ^23^, we took an alternative approach and instead used RNA sequencing as the ground truth for *ERBB2* expression levels. First, we separated the cohort into *ESR1* positive and negative groups using a cutoff of 6.2 TPM (**Supplementary Figure 9a**), which resulted in 98.3% (674/686 samples) of samples also being IHC positive. In the *ESR1* negative group, 78.5% (259/330) samples were IHC negative, while the majority (53.5%) of IHC positive samples contained low ER expression. Then we stratified the samples in both *ESR1* groups by using the interquartile range of *ERBB2* TPM (**Supplementary Figure 9a**), resulting in *ERBB2* positive (above 75^th^ percentile), *ERBB2* low (25^th^-75^th^ percentile), and *ERBB2* negative (below 25^th^ percentile).

As shown in **Extended Table 3**, differential expression analysis of these *ERRB2* expression subgroups revealed a minimal number of upregulated genes in *ERBB2* low relative to *ERBB2* negative, with the *ESR1* positive group showing the least upregulated genes (**Supplementary Figure 9b**). For the *ESR1* positive subgroup, only two genes were found to be upregulated in *ERBB2* low by both DeSeq2 and Wilcoxon, including *CRISPLD1*, a secretory protein that is part of the cysteine-rich secretory proteins (CAP) superfamily implicated in cancer and immunity, and the *UGT2A3* pseudogene. Expanding the analysis to upregulated DEGs by either DeSeq2 or Wilcoxon revealed several upregulated genes, including *KRT1*, *FLG*, *FLG2*, and *OLFM4*, with shared common functions as structural components of skin, cell adhesion processes, and protein binding ability. In the ESR1 negative subgroup, twelve genes were found to be upregulated in *ERBB2* low by both DeSeq2 and Wilcoxon. Most of these genes (*KLK10*, SLURP1, LINC02571, *AARD*, *SERHL2*, *SYT8*, *TNNI2*, and *ANGPT1*) were also found to be upregulated in the basal-like subtype relative to luminal and Her2 subtypes. Nevertheless, it is intriguing that the *ERBB2* low subgroup displayed overexpression of this markers relative to *ERBB2* negative, since the latter is more exemplary of a true basal-like phenotype. Notably ERBB2 was also upregulated in *ERBB2* low and some other genes located at the 17q12 locus (i.e., Her2 amplicon) were upregulated in the DeSeq2 (*PNMT*) and Wilcoxon (*GRB7*, *TBC1D3L*) analyses. In contrast, to the handful of upregulated genes, *ERBB2* low specimens, irrespective of *ESR1* status, showed broad downregulation of regulatory non-coding RNAs such as snoRNAs suggesting *ERBB2* low may have altered ribosomal RNA (rRNA) processing and post-transcriptional regulation (**Supplementary Figure 9b**, top right panels for ER positive and ER negative).

As expected, the genes that best differentiated *ERRB2* positive samples from *ERBB2* low and negative, irrespective of *ESR1* status, were located along the core of the Her2 amplicon (i.e., *ERBB2*, *PNMT*, *GRB7*, *TCAP*, *PGAP3*, *STARD3*, *MIEN1*) and are frequently co-amplified with *ERRB2*. We explored the Her2 amplicon further and found *ERRB2* low samples showed intermediate expression levels for many genes near the 17q12 locus when compared to either *ERRB2* positive or negative samples, a finding that was consistent across *ESR1* status (**Supplementary Figure 9c**). To see if expression of the Her2 amplicon could segregate *ERBB2* low from positive and negative samples, we calculated a Her2 amplicon module score and plotted each sample from highest to lowest score, which resulted in good separation of *ERBB2* positive, low, and negative samples (**Supplementary Figure 9d**). Altogether, these data suggest that *ERBB2* expression, select differentially expressed genes, and Her2 amplicon signatures could be used as an aid to stratify tumors into *ERBB2* positive, low, and negative, ultimately resolving the ambiguity of IHC for low Her2 expression.

### Laser capture microdissection (LCM) enables tumor-specific gene expression and accurate multigene profiling

Breast tumors display considerable intra-tumoral heterogeneity spanning histological, morphological/cellular, genetic, and molecular features, which has important implications for patient diagnosis, treatment, and prognosis. Particularly important for multi-gene assays (e.g., PAM50, OncotypeDX, etc.) is the tissue and cellular composition of the tumor because specimens with low tumor content (or tumor cellularity) can lead to spurious gene expression results. We found approximately 10.6% of tissues (n=115/1082) contained either histological (n=44/1082=4.3% for mixed histology and n=17/1082=1.7% for mixed invasive-DCIS/LCIS) or biomarker heterogeneity (n=16/1082=1.6% for all or none spatial expression and n=38/1082=3.8% high/low spatial expression; see Methods). To investigate the effects of tumor content and heterogeneity on gene expression, we selected a panel of 44 samples with variable tumor content and compared the transcriptome profiles of LCM with adjacent sections that did not undergo LCM (**Figure 5a**). LCM samples showed enrichment for each marker (*PGR*, *ESR1*, *ERBB2*, and *MKI67*), but only for samples that were classified as IHC-positive for the respective marker (**Figure 5b**). Genes that were classified as IHC-negative showed reduced expression when comparing LCM samples to matched undissected sections, except for *ERBB2*, which was either unaltered or enriched (**Figure 5c**). Interestingly, this observation may be related to the low levels of *ERBB2*/Her2 that may be present in specimens classified as Her2 negative (IHC 0) or Her2-low as defined by an IHC score of +1 or +2 without DNA amplification (for a review, see ^24–26^). LCM, compared to no LCM, also resulted in a broader dynamic range for all markers (**Figure 5d**), presumably because more sequencing reads are distributed to transcripts derived from cancerous tissues rather than normal, healthy epithelial, connective, and adipose tissues. In support of this, LCM enriched Cadherin-1 (*CDH1*) expression, a cell-type marker of breast glandular epithelial cells (i.e., tumor cell marker) and reduced the expression of Vimentin (*VIM*) and Platelet and Endothelial Cell Adhesion Molecule 1 (*PECAM1*), markers for fibroblasts and endothelial cells, respectively (**Figure 5e**).

**Figure 5.**
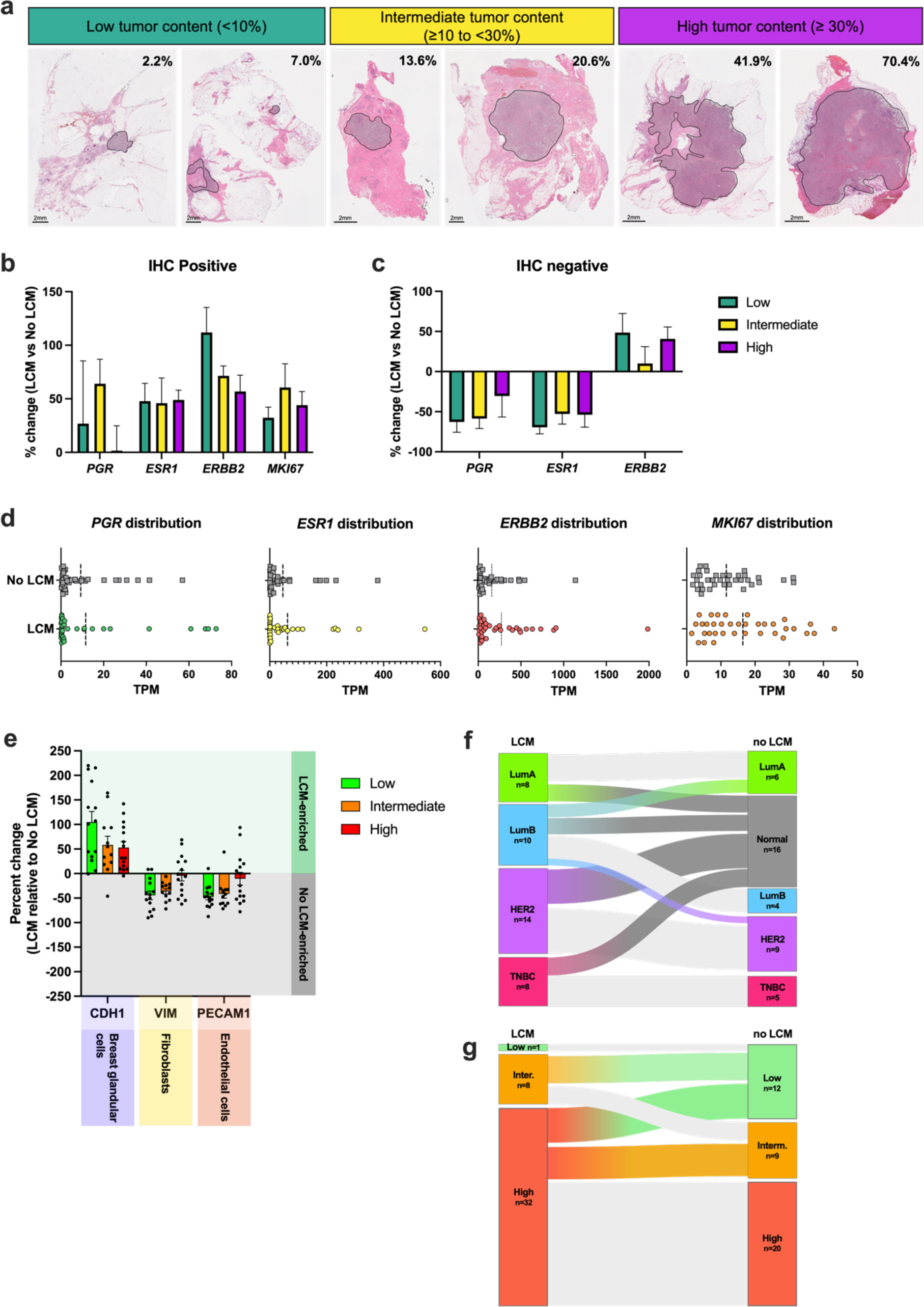
Comparison of laser capture microdissection with bulk processing on biomarker expression, molecular subtyping, and prognostic classifiers. (**a**) Photomicrographs depict examples of hematoxylin and eosin-stained resected tumor specimens with low, intermediate, or high tumor content represented in shaded annotations. Scale bars represents 2mm length. (**b,c**) Change in gene expression of *PGR*, *ESR1*, *ERBB2*, and *MKI67* in specimens that were classified as IHC positive (**b**) or IHC negative (**c**). (**d**) Dot plots show the dynamic range of gene expression for each biomarker in LCM vs no LCM matched samples. Dotted lines represent the median. € Expression of cell-type specific markers in LCM vs no LCM samples containing either low, intermediate, or high tumor content. (**f,g**) Sankey diagrams illustrate change in mFISHseq consensus subtypes (**f**) and in PAM50 risk of recurrence by subtype (ROR-S) classification (**g**) for LCM samples and their paired undissected scrolls.

We further explored the impact of LCM on molecular subtyping and prognostic risk classifiers as many of these procedures rely on bulk processing or tumor content thresholds to ensure reliable results. Molecular subtyping was particularly susceptible to the presence of non-tumor tissue with 61% (25/41) of AIMS and 32% (13/41) of both mFISHseq and PAM50 samples switching molecular subtype when comparing paired LCM and no LCM samples. This also resulted in 41% (17/41) samples switching their consensus subtype (**Figure 5f**). Expectedly, the most common consensus subtype change was to the normal-like subtype (15/41) and then two samples changed from LumB to LumA and one sample from LumB to Her2 (**Figure 5f**). Bulk processing also influenced the prognostic classifiers to a lesser extent with 41% (17/41) of ROR-S, 22% (9/41) of GGI, 15% (6/41) of GENE70, and 10% (4/41) of OncotypeDX samples switching prognostic risk groups. All samples that switched prognostic risk groups were classified as a lower risk group (e.g., high to intermediate or high to low), which could have profound implications for treatment since low-risk individuals may forego receiving potentially beneficial chemotherapy. For ROR-S, 12 patients (6 high and 6 intermediate risk by LCM) switched to the low prognostic risk group (**Figure 5g**) and these individuals would be incorrectly recommended to not receive chemotherapy. Overall, this highlights the importance of LCM in enriching tumor specific gene expression to provide accurate assessment of the four main breast cancer biomarkers and multigene testing. Methodologies, including commercial tests, that fail to adequately eliminate non-tumor elements may lead to erroneous gene expression, misclassification of patients, and dire consequences for prescribing inappropriate treatment regimens.

### Deployment of mFISHseq as a research-use only (RUO) test provides insights for tailored treatment

To demonstrate the clinical utility of mFISHseq, we conducted a RUO version of the assay on 26 patients. This version included our consensus subtyping and prognostic risk groups as well as 40 genes and 28 gene signatures spanning multiple cancer-related pathways relevant for treatment and prognosis (**Figure 6a**), including 20 ADC antigen targets and 5 payload-related targets (**Figure 6b,c**). Genes and gene signatures were compared with either all patient samples from our retrospective clinical validation or patient samples restricted to the relevant IHC-surrogate subtype to assign a percentile ranking score that was then categorized in tertiles (high, intermediate, low). These patients spanned neoadjuvant, adjuvant, and advanced/metastatic clinical settings and were representative of all molecular subtypes, especially basal-like/TNBC (**Figure 6d**). The most frequently recommended therapies by mFISHseq are dominated by more novel, targeted therapies such as ADCs, PARP inhibitors, and immunotherapies (**Figure 6e**). Below we outline several cases and a detailed report of each patient can be found at https://multiplex8.com/medical-professional (Note that the following patient ID numbers listed below do not reveal any identifying information and are not known to anyone outside of the authors of this manuscript).

**Figure 6.**
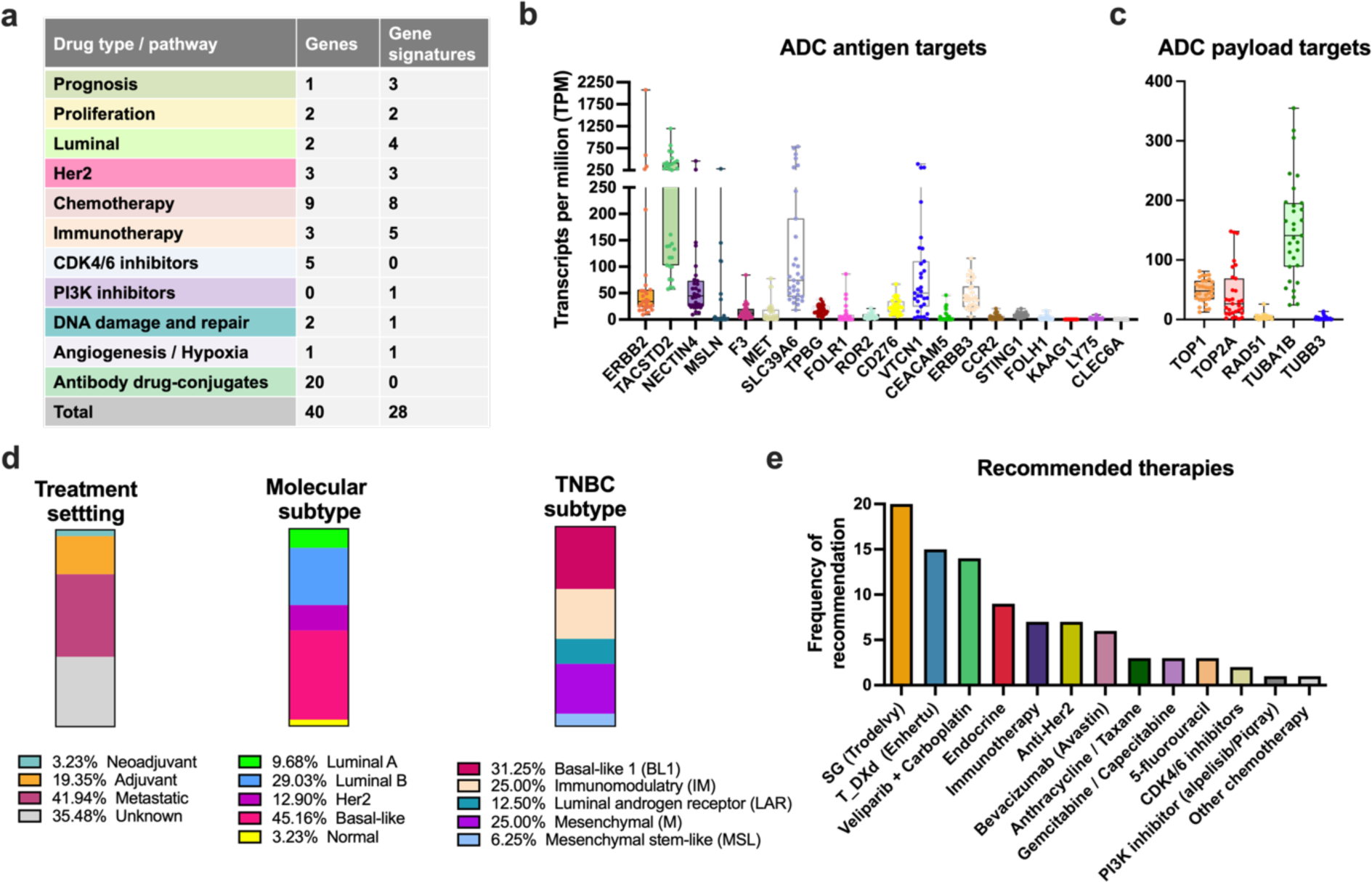
Deployment of mFISHseq as a research-use only test. (a) Table outlining the number of genes and gene signatures and their relevant drug targets/pathways that were used for clinical testing. **(b,c)** Expression of 20 ADC antigen targets (**b**) and targets relevant to payloads for topoisomerase and microtubule inhibitors (**c**) in 26 patients. (**d**) Proportion of 26 patients according to treatment setting (left bar), molecular subtype (middle bar), and TNBC subtype (right bar). (**e**) Frequency of therapies that were recommended as the top 3 according to expression of genes and gene signatures tailored to each of the 26 patients.

Patient 1 had TNBC and received a taxane/carboplatin regimen in the neoadjuvant setting without effect leading to a mastectomy. The resected tumor was classified as invasive ductal carcinoma (pT3, G3, pN1) and ER/PR/HER2− with 60% Ki67. After surgery, the patient received capecitabine and then had a locoregional relapse. We conducted mFISHseq on the resected tumor and the histopathology revealed two distinct regions of the tumor that were characterized either with or without infiltrating lymphocytes. We dissected both regions (Sample A and B) and mFISHseq confirmed the IHC results (*PGR*/*ESR1*/*ERBB2*− and *MKI67* high) and basal-like intrinsic subtype for both ROIs; however, the two regions differed in their TNBC subtypes with Sample A and B being classified as mesenchymal (M) and immunomodulatory (IM), respectively. To identify treatment predictive signatures that were common between the two ROIs, we investigated a tailored list of genes and gene signatures relevant for metastatic TNBC, revealing that both ROIs contained high scores for an angiogenesis/hypoxia/inflammation signature that may predict response to bevacizumab (Avastin) as well as high expression for *TACSTD2* and *TOP1* (i.e., antigen and payload targets for sacituzumab govitecan (Trodelvy). This provided two viable treatment options in the first and second lines for metastatic TNBC. Notably, this evidence concurred with follow up data from this patient went into complete remission following bevacizumab treatment.

Another two patients, Patient 15 and Patient 19, received T-DM1, thus allowing us to test our T-DM1_pred signature. Patient 15 is a 46-50-year-old female with invasive ductal carcinoma (pT1c, G2) that was ER/PR/HER2+ with high Ki67. She received doxorubicin/cyclophosphamide plus docetaxel followed by dual Her-2 blockade with trastuzumab/pertuzumab as neoadjuvant treatment but had disease progression prior to surgery. Currently, she is receiving adjuvant T-DM1 and awaiting follow-up. Patient 19 is a 70-75-year-old female with invasive ductal carcinoma (pT4b, G2, node positive) that was ER+/PR−/HER2+ (IHC 2+ with positive amplification by DNA-FISH) with 15% Ki67. She received T-DM1 in the metastatic setting and is currently in stable condition. For Patient 15, mFISHseq agreed with IHC for all markers, classified the sample as luminal B, and showed high expression of *ERBB2* and *ERBB2*-signatures (Her2 amplicon_MDX and Module7_ERBB2), low-high expression of Her2 resistance markers, and a moderate T-DM1_pred score (score = 0.29; 57^th^ percentile) that was above the threshold for predicting response to T-DM1. Patient 19 had two heterogeneous ROIs based on high (Sample A) and low (Sample B) *ERBB2* expression that were dissected with mFISHseq. The mFISHseq results for both ROIs agreed with IHC results, except for Sample B, which was classified as *ERBB2* low and had lower *ERBB2*-signatures (Her2 amplicon_MDX and Module7_ERBB2). Similarly, only Sample A had a T-DM1_pred score (score = 0.03; 48^th^ percentile) that was above the threshold for predicting response to T-DM1. This anecdotal evidence is supported by the stable disease of this patient following T-DM1 treatment in the metastatic setting.

Patient 3 is a female patient with TNBC in the adjuvant setting who has received standard-of-care chemotherapy (AC, doxorubicin + cyclophosphamide). The mFISHseq test classified this patient as *ESR1*/*PGR*/*ERBB2*−/*MKI67*+, basal-like consensus subtype, and IM TNBC subtype, which was also supported by high levels of immune-related genes (*PDCD1*, *CD274*, *CTLA4*) and gene signatures (e.g., Chemokine12, Dendritic_cells, etc.). This suggests treatment with immune checkpoint inhibitors like pembrolizumab (Keytruda) or atezolizumab (Tecentriq) may be beneficial in addition to standard chemotherapy. This patient also had high levels of *TACSTD2* and *TOP1*, the antigen and payload targets for the anti-Trop-2 ADC, sacituzumab govitecan (Trodelvy). This highlights the potential of mFISHseq to predict potentially beneficial therapies in subsequent treatment lines, which may save valuable time in selecting new treatments after a patient becomes refractory.

As noted earlier, the consensus prognostic categories of mFISHseq may have utility in identifying patients with low (ultra-low) risk who can safely forego adjuvant chemotherapy and this potential is demonstrated in the report for Patient 23. This female patient (age 46-50) has a clinically low risk invasive ductal carcinoma (pT1b, G2, pN0) that is ER/PR+, Her2− (low, 2+), and 15% Ki67. In addition to agreeing with the IHC results and classifying the patient as luminal A, mFISHseq found low risk classifications in four multiparameter tests (OncotypeDX, GENE70, ROR-S, and GGI) and clinical risk, suggesting this patient is classified as ultra-low and can be spared at least potentially toxic chemotherapy without having a detrimental outcome.

Overall, the RUO version of mFISHseq (called Multiplex8+) provided unique insights for each patient that could identify potential treatments (including subsequent lines of therapy) and helped to explain why prior treatments performed poorly. Like our retrospective study, there was exceptional concordance between IHC results and mFISHseq with agreement in 93% (n=87/94) of cases for all four biomarkers, with Her2 IHC and *ERBB2* mFISHseq showing perfect agreement. All discordant results were observed for *PGR* (n=3) and *ESR1* (n=4). In two patients (#7 and #12), both ER and PR were in gray zone areas for IHC (i.e., ER/PR low ≤10%) and mFISHseq classified them as negative. Notably, both patients were classified as basal-like by consensus molecular subtyping, mesenchymal (M) TNBC subtype, and had high expression of cancer hallmark pathways like proliferation, immune, and/or angiogenesis suggesting the mFISHseq results reflected the underlying tumor biology better than the IHC markers.

## Discussion

The mFISHseq test utilizes two orthogonal methods, RNA-FISH and RNA-SEQ, to characterize breast tumor biology. Simultaneously visualizing gene expression of the four main breast cancer biomarkers in a multiplexed and multicolor RNA-FISH reaction allows users to identify ROIs based on cellular phenotypes and tumor heterogeneity and then precisely capture these ROIs using LCM for downstream spatially resolved transcriptomics. Total RNA sequencing facilitates transcriptome-wide expression profiling to classify the molecular subtype of breast cancer and quantify predictive and prognostic gene signatures.

In this retrospective validation cohort of 1,082 FFPE breast cancer specimens, mFISHseq showed high concordance with biomarkers assessed by IHC, a finding that is consistent with other reports ^27,28^ and highlights the analytical validity of the assay; however, the concordance varied depending on biomarker IHC expression level. This is consistent with reports showing that IHC concordance among pathologists in these low ranges is poor due to the limitations of IHC (pre-analytical factors, variable antibody specificity/sensitivity, subjective quantification, etc.). For example, a recent study showed that a group of 18 pathologists had only 26% concordance between HER2 specimens containing H-scores of 0 and +1, which could result in misclassification of patients and consequently eligibility for the new anti-HER2 drug conjugate trastuzumab deruxtecan (Enhertu) ^23^. Thus, novel quantitative assays that can accurately define Her2 expression are critical to identify patients eligible for emerging anti-Her2 drug conjugates.

Many researchers have approached this challenge by exploring biological and clinicopathological differences of Her2-low tumors albeit with disparate results. To address this, we surmised that RNA sequencing expression data of *ERBB2* may provide a more suitable ground truth compared to IHC, which is known to suffer from poor concordance when classifying Her2-low (IHC +1/+2 unamplified by DNA ISH) and negative (IHC 0) specimens ^23^. Therefore, we used our RNA sequencing data to stratify *ERBB2* expression in respect to ER status and conducted robust differential expression analyses using two approaches to mitigate false positive results. Few upregulated genes were observed in *ERBB2* low, relative to *ERBB2* negative. In contrast, downregulation of genes accounted for most of the differences between *ERBB2* low and negative, with *ERBB2*-low samples showing reduced expression of a broad panel of snoRNAs and other regulatory RNAs, irrespective of ER status. This is a novel finding with important implications for Her2-low given the broad regulatory roles of snoRNA in processing of rRNA and post-transcriptional regulation together with evidence of their dysregulation in breast cancer ^29–32^. This suggests that rather than being a distinct biological subtype akin to luminal A/B, Her2-enriched, and basal-like/TNBC, Her2-low specimens are mainly characterized by subtle changes in rRNA processing and post-transcriptional regulatory programs on top of their main molecular background (e.g., luminal vs basal). We also demonstrate several novel approaches to quantify Her2 in low expression ranges by using RNA expression assays like RNA-FISH and RNA-seq to characterize *ERBB2* levels as well as genes located near the 17q12 Her2 amplicon locus. These approaches could supplement current IHC diagnostics to provide additional information about Her2 expression in gray zone cases.

Although molecular subtyping can provide clinically relevant information, several reports have documented problems with discordance at the single-sample level ^7,8^, which has often been attributed to research based versions of the test as well as differences in data processing. However, in the OPTIMA study, the Prosigna, Blueprint, and MammaTyper test showed marked disagreement in 40.7% of patients resulting in only moderate concordance between each test (κ range = 0.39-0.55), despite each test being conducted by the respective vendors ^9^. The discordance and irreproducibility have impeded widespread adoption of multigene subtyping tests in clinical practice even though there is broad acknowledgment of their potential to yield clinical insights. Our findings echoed these reports as we observed only moderate concordance between three multigene classifiers and the IHC surrogate subtypes as well as substantial concordance among the three multigene classifiers, with the most and least discordance in luminal A/B subtypes and in basal-like/Her2 subtypes, respectively. Each multigene subtyping tool uniquely classified a subset of samples that no other classifier agreed with, and these samples generally showed evidence of misclassification in survival, subtype stability, and prognostic risk. Our consensus subtyping, based on a simple voting scheme of the three multigene classifiers (mFISHseq, AIMS, and PAM50) effectively alleviated this misclassification by reclassifying approximately 30% of these samples into subtypes that better corresponded to their overall survival and prognostic risk. Thus, consensus subtyping resolves the propensity for individual molecular subtyping classifiers to assign subtypes to single samples with unacceptable levels of discordance.

Several groups have attempted to address the discordance among multiparameter tests at the single sample level usually be addressing methodological challenges such as composition of a reference/training set and data normalization/processing ^33–36^. One approach constructed a consensus training set of samples that were unanimously subtyped by multiple classifiers and then used this to train additional single-sample predictors that then showed near perfect concordance (median κ>0.8) ^37^. Notably, they also demonstrate that changes in the gene list have a greater influence on intra- and inter-predictor concordance than the training set and type of predictor. This finding supports our consensus subtyping approach that leverages total RNA transcriptomics to simultaneously derive subtype predictions by combining several multigene classifiers and consequently several different gene lists. Thus, consensus subtyping addresses the main culprit for discordance, which is the use of disparate gene lists to derive a categorical subtype that may have subtle biological differences based on the assay and gene list used. This critical point is elaborated on in a recent perspective where Schettini, Prat, and colleagues argue that the construct of a molecular subtype is not necessarily interchangeable, since each multiparameter assay interrogates different biomarkers with distinct methodologies^38^.

Like consensus subtyping, we found that combining information from 5 different prognostic classifiers into a consensus prognostic classification system of high, low, and ultra-low risk groups provided more accurate results for cases with discordance in at least one classifier. Moreover, the ultra-low risk patients (i.e., those with agreement in all 5 classifiers with node negative and PR+ status) had excellent PFS and OS out to 15 years. With this consensus prognostic classification system, high risk individuals showed the poorest OS and more relapses within 5 and 10 years and benefitted the most from chemoendocrine therapy. Low risk patients had better OS, more relapses after 10 years, and showed no benefit from chemotherapy in addition to endocrine therapy. Notably, many of these patients received only 5 years of endocrine therapy and may constitute a group of patients that could benefit from extended endocrine therapy for 10 years. The ultra-low risk group contained about 10% of patients that had low probability of relapse and excellent survival. This group may overlap with ultra-low risk categories identified by other studies, which is roughly 10-25% of all patients depending on screening and may represent a group of patients with indolent tumors that need only surgery and no adjuvant systemic therapy at all.

The substantial inter-predictor concordance we observed without consensus subtyping was interestingly higher than prior reports ^7–10^. One potential explanation for this higher concordance is the use of laser capture microdissection to precisely capture tumor-enriched samples for downstream RNA-SEQ. Most other studies use bulk specimen processing (or microdissection), which not only loses the spatial information about biomarker expression but may introduce erroneous gene expression from healthy stroma/epithelia, ductal/lobular carcinoma in situ, and other non-tumor tissue, which can lead to RNA-based gene expression profiling tests to misclassify breast cancer samples into molecular subtypes and prognostic risk groups ^8,39–43^. For example, the “normal-like” subtype is simply a byproduct of insufficient tumor content/cellularity. Similarly, the six TNBC subtypes ^21^ were refined to four when LCM was used to demonstrate that the immunomodulatory and mesenchymal stem-like subtypes result from elevated infiltrating lymphocytes and tumor-associated mesenchymal cells, respectively ^20^. In our comparison of paired sections that underwent LCM or bulk processing, we found that bulk processing resulted in substantial changes in gene expression and assignment to molecular subtypes and prognostic risk groups, which generally resulted in downscaling to less aggressive subtypes (e.g., normal-like) and risk groups. This has important implications for treatment as some patients, especially those in gray zones (e.g., bordering high/low risk thresholds) may be misclassified and consequently inappropriately treated. This stresses the importance of incorporating laser capture microdissection into the mFISHseq workflow to minimize contamination from non-tumor elements and obtain spatially defined, tumor-enriched cell populations, ultimately facilitating precise and accurate biomarker quantification and multiparameter testing.

Recent evidence from the clinical trials for the recently approved ADCs, trastuzumab deruxtecan (Enhertu) ^22^ and sacituzumab govitecan (Trodelvy) ^44,45^, suggest that protein expression of the antigen target is insufficient to predict treatment response, highlighting in unmet need to identify novel biomarkers that can stratify patients into those most likely to response. In this study, we conduct one of the most comprehensive analyses of ADC targets and ADC processing-related genes and gene signatures by characterizing their expression patterns and association with survival in over 1000 patient samples. The results portray unique subtype-specific expression patterns that may be relevant for stratifying patients into ADC-responsive subgroups. We validated this hypothesis using data from the trastuzumab emtansine arm of the I-SPY2 trial, where we identified a gene signature containing 19 features related to the ADC target (*ERBB2*), receptor endocytosis, lysosome function, and microtubule targets of the maytansine payload. Moreover, this signature outperformed *ERBB2* expression alone in a multivariate logistic regression, thus supporting the notion that combining ADC processing features into a predictive model has clinical utility for patient selection. Notably, both ERBB2 mRNA and our T-DM1_pred had substantially more predictive utility than Her2 IHC, since 42.3% of patients (n=22/52) did not responds despite being Her2 IHC positive. An important next step will be to validate this signature as a prespecified, qualified biomarker in a larger external cohort. Overall, these data create a foundation and roadmap for identifying subgroups of patients that are more likely to respond to an ADC, and each signature can be tailored to key features of the ADC like antigen and payload target (topoisomerase, microtubule, or DNA), cleavable (enzyme or acid labile) or noncleavable (lysosome processing) linker, and mechanisms of resistance (ABC transporters, glucuronidation enzymes).

A limitation of the current study is the retrospective design, which is susceptible to confounding and biases introduced by patient selection, misclassification, and recall. We attempted to mitigate this limitation prior to initiating the study through the following measures: 1) drafting a study protocol with a sample size/power analysis and prespecified data collection and statistical analysis plan; and 2) sourcing archived tissues from reputable biobanks (Biobank Graz and PATH Biobank) according to BRISQ guidelines. During the study, we also mitigated technical errors and other biases that could arise by processing many batches of samples by 1) pseudo randomizing and stratifying batches to ensure proportional representation of IHC surrogate subtypes in each batch; 2) ensuring research personnel were blind to specimen clinical information; and 3) implementing batch processing controls (ERCC spike-in and positive controls) for RNA-SEQ library preparation. These measures together with the excellent analytical validity relative to IHC and transcriptomic findings that are congruent with prior work (e.g., molecular subtyping and gene signature) and supported by outcome data further highlight the robustness and veracity of the results.

In this retrospective validation of mFISHseq on a cohort of 1,082 breast tumors, we provide analytical validity that mFISHseq can be used with high confidence to assess the expression of the four main breast cancer biomarkers currently used in IHC testing. Both methods can be used to cross-validate one another to mitigate false positives/negatives due to preanalytical and technical factors, and consequently making strides to correctly classify equivocal cases. By using laser-capture microdissection to collect spatially defined, tumor-enriched samples, mFISHseq provides tumor-specific gene expression that can be used to derive insights about the tumor microenvironment, molecular subtype, and tumor biology that may predict treatment response and outcome. We further demonstrated this clinical utility on 26 patients who received an RUO version of the test, which we named Multiplex8+ (patient reports are located at www.multiplex8.com (in the medical professional section).

## Methods

### Study Design

We conducted a retrospective clinical validation of our mFISHseq BCa diagnostic test using 1,082 archived FFPE BCa samples collected from two European biobanks (Biobank Graz, PATH Biobank), one hospital (Malaga, Spain), and two commercial companies (AMS Bio and Precision for Medicine). Clinicopathological information associated with each sample (age, receptor histological status, tumor grade, therapy history, survival data, etc.) was accrued in collaboration with the biobanks and follows the Biospecimen Reporting for Improved Study Quality (BRISQ) criteria and used to perform association analyses with molecular data. Inclusion criteria for the study consisted of females with histologically confirmed invasive BCa, availability of anonymized data regarding pathological diagnosis (IHC status, TNM staging), therapy (hormone/targeted/chemo- or radiotherapy), and survival (progression-free survival, overall survival), as well as signed and dated informed consent. The only exclusion criteria were pre-existing conditions or concurrent diagnosis of a cancer other than breast cancer or other disease that may influence the interpretation of the study results. The Ethics Committee of the Bratislava Self-Governing Region gave ethical approval for this work (Ref. No. 05320/2020/HF). In addition, the Ethics Commission of the Medical University of Graz on behalf of Biobank Graz gave ethical approval for this work (No. 34-354 ex 21/21, 1158-2022).

Patient specimens were processed in batches (see **Supplementary Methods**) using a stratified randomization approach to ensure that each batch contains a representative sampling of the IHC-surrogate subtypes (i.e., luminal A, luminal B, HER2+, TNBC). Researchers who processed batches and conducted the data processing and analysis were blind to IHC biomarker status (e.g., ER, PR, HER2, and KI67) and other clinical information.

To assess the analytical validity of mFISHseq, the dataset was divided into a training and test set (70:30 split) using a stratified randomization approach to ensure similar proportions of positive and negative biomarkers (as defined by IHC) and sufficient patient outcomes. Other analyses like consensus subtyping and characterization of genes/gene signatures utilized the full dataset.

### Tissue processing and H&E staining

We obtained at least eight 5 μm sections from FFPE BCa specimens using a Leica Histocore Multicut. Adjacent sections were collected in the following order: 1 section on a glass slide for H&E, 1 section on a functionalized PEN membrane slide for LCM, 1 section on a glass slide for RNA-FISH, and 1 section taken as a scroll and frozen at –20 °C for later RNA extraction and RNA sequencing (see **Supplementary Methods**). To ensure proper identification of invasive breast cancer, we stained one section using H&E, cover slipped, and then obtained a whole-slide scan. The resulting image was annotated by a trained researcher to identify the invasive breast cancer component for later microdissection. If necessary, a board-certified pathologist either annotated or reviewed challenging cases. Importantly, the H&E-stained section was adjacent to the PEN membrane slide used for laser capture microdissection to ensure comparable anatomical morphology between the slide that was annotated and the slide that was microdissected.

### Multiplexed RNA-FISH

For RNA-FISH we used the Advanced Cell Diagnostics RNAscope™ Multiplex Fluorescent V2 Assay to detect *PGR*, *ESR1*, *ERBB2*, and *MKI67* according to the manufacturer’s instructions. These markers were detected using Akoya Opal 690, 620, 520, and 570 fluorophores, respectively. Visualization of these markers allowed us to capture the heterogeneity of the tumor tissue and isolate key regions of interest using LCM to obtain tumor-specific regions of interest, while eliminating otherwise healthy tissue, stroma, and adipose cells that may “mask” true gene expression differences.

### Whole-slide imaging, image annotation, and image analysis

Following H&E staining and RNA-FISH, we used the Akoya Vectra Polaris to obtain brightfield and fluorescent whole slide scans (20x objective) that could be further analyzed and annotated for microdissection. The H&E whole slide scans were annotated by a trained researcher using the open-source program QuPath (https://qupath.github.io/). Annotations were color-coded to identify invasive breast cancer (segregated by histological subtype if more than one is present in a specimen), ductal carcinoma in situ (DCIS), and healthy/normal tissue. If necessary, a board-certified pathologist either annotated or reviewed challenging cases. The RNA-FISH whole slide scans were annotated by a trained researcher using Akoya’s Phenochart software (Vectra Polaris). The RNA-FISH annotation consisted of a qualitative overview of the intensity and distribution of fluorescent signals from ER, PR, HER2, and KI67. Based on the annotated H&E and RNA-FISH images, specific regions of interest were selected for LCM with an emphasis on regions that displayed the expression of biomarkers of interest (e.g., hotspots), areas identified as invasive by a trained researcher/pathologist, and the margins of invasive tumors. Moreover, in the case of specimens that displayed histologic or biomarker expression heterogeneity in the form of different molecular expression patterns (e.g., ER/PR+ and HER2-regions versus ER/PR- and HER2+ regions) or histological subtypes (e.g., invasive ductal vs invasive lobular carcinoma) distinct regions of interest were annotated and separately subjected to LCM and downstream analyses (**Supplementary Methods**).

For RNA-FISH image analysis, regions of interest that were dissected by laser capture microdissection were stamped on the digital whole slide scans for further processing of biomarker signals using Akoya’s InForm software (**Supplementary figure 4**). At least 1-3 stamped regions, depending on the size of the area dissected by LCM, were analyzed (at 20x objective). The analysis consisted of the following steps: 1) spectral unmixing and autofluorescence isolation using a synthetic spectral library; 2) using machine learning algorithms to segment the tissue into different regions (tumor versus stroma) as well as to segment individual cells into nuclear and cytoplasmic components; and 3) scoring the expression of each biomarker. The average fluorescence intensity for each marker was assessed specifically in the tumor segment of the image and the researcher conducting the analysis was blinded to the known IHC results and the clinicopathological data.

### Laser capture microdissection

We followed established protocols from Leica for conducting LCM in a manner that maintained RNA integrity. This included conducting a rapid, cresyl violet stain, limiting dissection times to under 1 hour per sample, and taking precautionary measures to ensure RNA integrity. Regions selected for dissection were identified by comparing the annotated H&E and RNA-FISH images with the adjacent cresyl violet stained section. We aimed to dissect approximately 10-20 mm^2^ of tissue per sample to ensure an adequate amount of material for RNA extraction. For samples with less tumor area, we conducted LCM on multiple PEN membrane slides to obtain sufficient tissue.

### RNA isolation and quality control

The Macherey Nagel NucleoSpin totalRNA FFPE XS kit was used for RNA isolation (see **Supplementary Methods**). After RNA isolation, RNA quantity was measured using the Qubit RNA HS (High Sensitivity) Assay Kit with a Qubit 4 Fluorometer and RNA quality using the Agilent High Sensitivity RNA ScreenTape with an Agilent 4150 TapeStation. The DV_200_ value of the sample (i.e., the percentage of fragments ≥ 200 bases in length) was calculated as recommended by Illumina. Samples with DV_200_ values > 15% were considered as viable samples for library preparation.

### RNA library preparation and sequencing

We used the Takara SMARTer Stranded Total RNA-Seq Kit v3 - Pico Input Mammalian kit to prepare total RNA-SEQ libraries following the manufacturer’s instructions. To control for batch library preparation effects, we included a single natural positive control sample in each library preparation batch and a synthetic spike-in control in each sample (see **Supplementary Methods**). Following library preparation, the quantity and fragment size range of the library were assessed using both the Qubit dsDNA HS kit (Qubit 4 Fluorometer) and the Agilent High Sensitivity DNA ScreenTape kit (Agilent 4150 TapeStation). Successfully prepared libraries contained sufficient library (≥ 4ng/μl) to pool on an Illumina NovaSeq 6000 sequencing instrument and fragment range spanning 200 – 1,000 bp, with a local maximum ∼250 – 350 bp. Individual sequencing libraries were pooled and sequenced on an Illumina NovaSeq 6000 using SP, S1, S2, or S4 flow cells depending on pool size). Pooled libraries were spiked with 10% PhiX as recommended by both Illumina and Takara for low-complexity libraries sequenced on patterned flow cells. Paired-end sequencing (2 x 100 bp) was conducted with the aim of obtaining approximately 100 million reads per sample. The bioinformatics pipeline for RNA sequencing is described in **Supplementary Methods**).

### Statistical Methods

All statistical tests were conducted using GraphPad Prism 9 and R studio packages. Unless otherwise stated, the level of significance was set at P < 0.05 for both adjusted (q-value) and unadjusted P-values. For determining the significance of continuous variables, paired or unpaired t-tests were used for normally distributed data for two groups, while linear mixed models or ANOVA were used for normal data for three or more groups. Non-normal data for two groups were analyzed using Mann-Whitney *U* tests (independent samples), while non-normal data for three or more groups were analyzed using Kruskal-Wallis tests. Appropriate corrections for multiple comparisons were conducted using Tukey’s test (parametric), Dunn’s test (non-parametric), or Benjamini-Hochberg (or Benjamini, Krieger, and Yekutieli) FDR procedures (non-parametric) to adjust P values. ROC and precision-recall curves were constructed using either GraphPad Prism 9 or the R package pROC 1.3.1. Diagnostic performance metrics were calculated using the R package caret 6.0-94.

#### Cohort descriptive statistics

– Standard descriptive statistics (presented as either median or mean ± standard error of the mean (SEM)) were used to summarize sample characteristics (e.g., data for biospecimen information, demographics, pathological, therapy type, survival) for all specimens in the study and were segregated by source and IHC-surrogate subtype.

#### RNA-FISH

– For each RNA-FISH target (*ESR1*, *PGR*, *ERBB2*, *MKI67*), fluorescent intensity (normalized to exposure time and segregated into the cytoplasm, nucleus, and total cell) was assessed. Standard descriptive statistics and violin plots (presented as either Median or Mean ± SEM or interquartile range) were used to summarize results. To compare between RNA-FISH and known IHC results within individual specimens, continuous data (e.g., fluorescent intensity), were analyzed using Pearson correlations for continuous data (e.g., fluorescent intensity) and Spearman’s correlations for ordinal data.

#### Survival / Outcome analyses

– Kaplan-Meier analysis and/or Cox Proportional Hazard models were used to quantify associations made between specific dependent variables and/or genes and gene signature predictors with known clinical outcome data (overall survival, progression-free survival). Both univariate and multivariate analyses with clinical parameters (tumor size and node status) were conducted using the R package survival (v3.5-7).

## Data Availability

The data that support the findings of this study are available within the paper and its supplementary information files. Reports from the 26 patients that underwent a research use only version of the diagnostic test (called Multiplex8+) can be found at https://www.multiplex8.com/medical-professional. The underlying data, custom code and scripts used for bioinformatic analyses, and access to the full dataset for research and/or commercial use may also be obtained upon a reasonable request from the lead corresponding author (P.&Ccaron.).

## Acknowledgements

We gratefully acknowledge Biobank Graz and PATH Biobank for providing the FFPE breast cancer specimens and associated clinicopathological data; Jeff Palatini, Dorota Adamska, Krzysztof Goryca, and other staff at the Centre for New Technologies, University of Warsaw, Genomics Core Facility for providing first-class sequencing and bioinformatic services; Dr. Karol Kajo, a board-certified pathologist, at the St. Elizabeth Cancer Institute Hospital in Bratislava for assistance in annotating digital whole slide images of H&E-stained tissues. This project was supported by the European Union’s Horizon 2020 research and innovation programme under an EIC Accelerator grant (agreement No 946693) awarded to MultiplexDX s.r.o. (Pavol Cekan as PI). FP is funded in part by an NIH/NCI P50 CA24779 01 grant. The funding sources had no role in the design and conduct of the study, interpretation of the data, writing of the manuscript, and decision to submit the manuscript for publication.

## Author contributions

E.D.P, B.H., N.Val., N.B., S.B., V.Man., T.T., F.P., and P.Č. conceptualized and planned the study. B.H., D.G., S.G., H.I., T.O., J.B., K.B., Z.K., and N.Voj., sectioned FFPE tissues, conducted histological staining and RNA-FISH, annotated H&E images, quantified RNA-FISH images, and performed laser capture microdissection. N.Val., N.B., S.B., D.D., M.K., D.L., V.Mam., and V.Man., extracted RNA, prepared RNA-SEQ libraries, conducted RNA/cDNA quality control, and pooled libraries for sequencing. N.Val., S.G., H.I., M.R., and P.M. conducted bioinformatic analyses. E.D.P., B.H., N.Val., N.B., D.G., S.G., H.I., T.O., S.B., J.B., K.B., D.D., Z.K., M.K., D.L., V.Mam., V.Man., N.Voj., M.R., I.C.M, I.A., P.M., T.T., F.P., and P.Č. verified the underlying data, analyzed, and interpreted the data, prepared figures, and wrote the manuscript. All authors had full access to the data, provided critical comments and feedback on the manuscript, and accepted responsibility to submit the manuscript for publication.

## Ethics declaration

The Ethics Committee of the Bratislava Self-Governing Region gave ethical approval for this work (Ref. No. 05320/2020/HF). In addition, the Ethics Commission of the Medical University of Graz on behalf of Biobank Graz gave ethical approval for this work (No. 34-354 ex 21/21, 1158-2022).

## Code and Data Availability

The data that support the findings of this study are available within the paper and its supplementary information files. Reports from the 26 patients that underwent a research use only version of the diagnostic test (called Multiplex8+) can be found at https://www.multiplex8.com/medical-professional. The underlying data, custom code and scripts used for bioinformatic analyses, and access to the full dataset for research and/or commercial use may also be obtained upon a reasonable request from the lead corresponding author (P.Č.).

## Extended Data

**Extended Figure 1.**
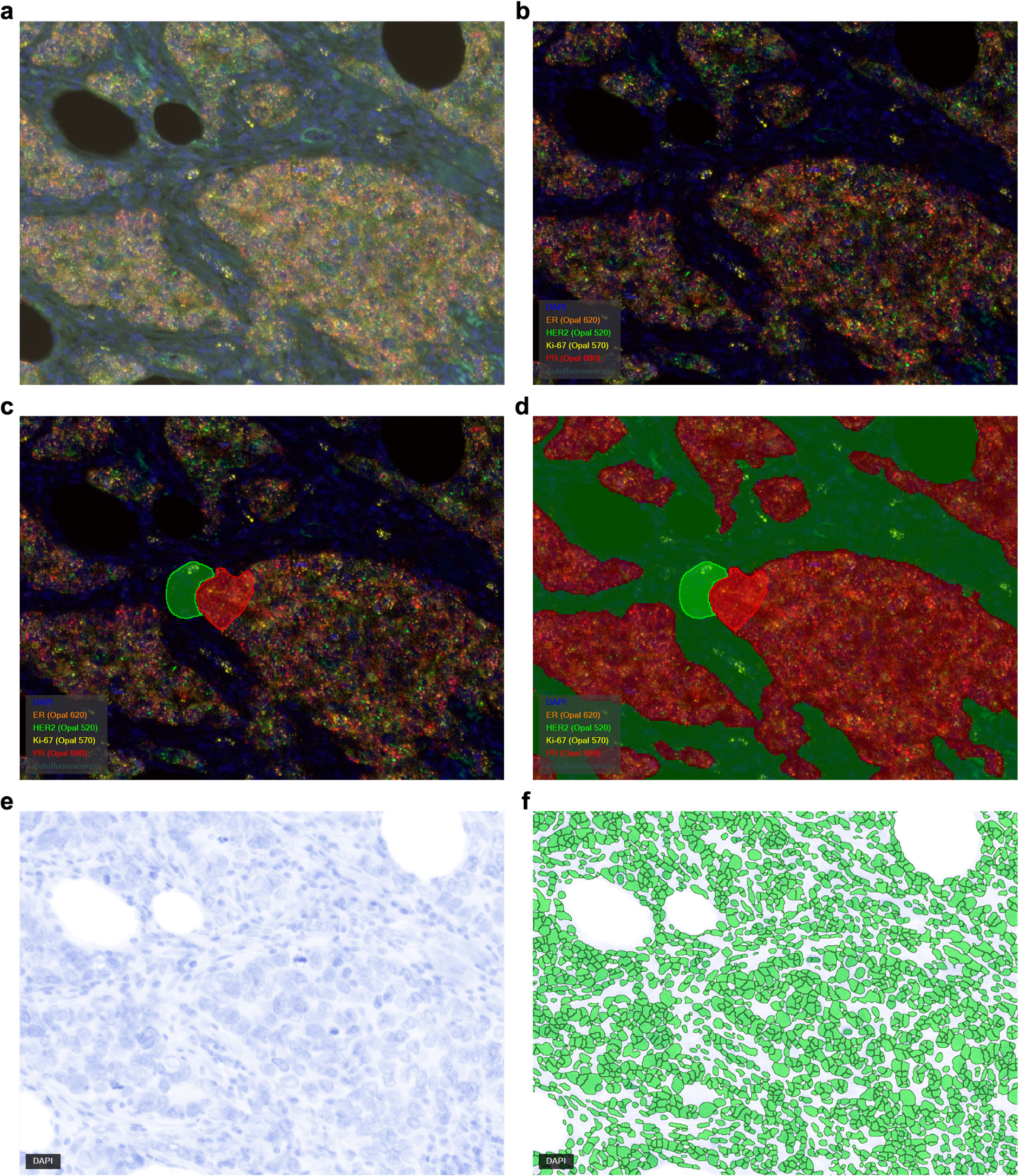
RNA-FISH image analysis pipeline using digital pathology/machine learning. Representative images depicting the RNA-FISH image analysis pipeline for tissue and cell segmentation. The tissue was stained for DAPI, ERBB2 (Opal520), MKI67 (Opal570), ESR1 (Opal620), and PGR (Opal690) and scanned at 20x. **(a)** The image analysis workflow started with raw images of multiple merged fluorescent channels. **(b)** The same image was split into individual separate fluorescent channels - DAPI, Opal520, Opal570, Opal620, Opal690, and autofluorescence, according to the spectral library for each fluorescent probe, and the autofluorescence was isolated from the image. **(c)** Using a machine learning algorithm, the tissue was segmented based on the selected training regions (red = tumor; green = stroma) with the training considering the fluorescent signal intensity of DAPI, ERBB2, MKI67, ESR1, and PGR. **(d)** Then the trained algorithm was applied to the whole image. **(e)** Individual cells were identified by the shape and intensity of DAPI staining, **(f)** then segmented, and fluorescent intensity values were normalized to exposure time and obtained for both tumor and stroma.

**Extended Figure 2.**
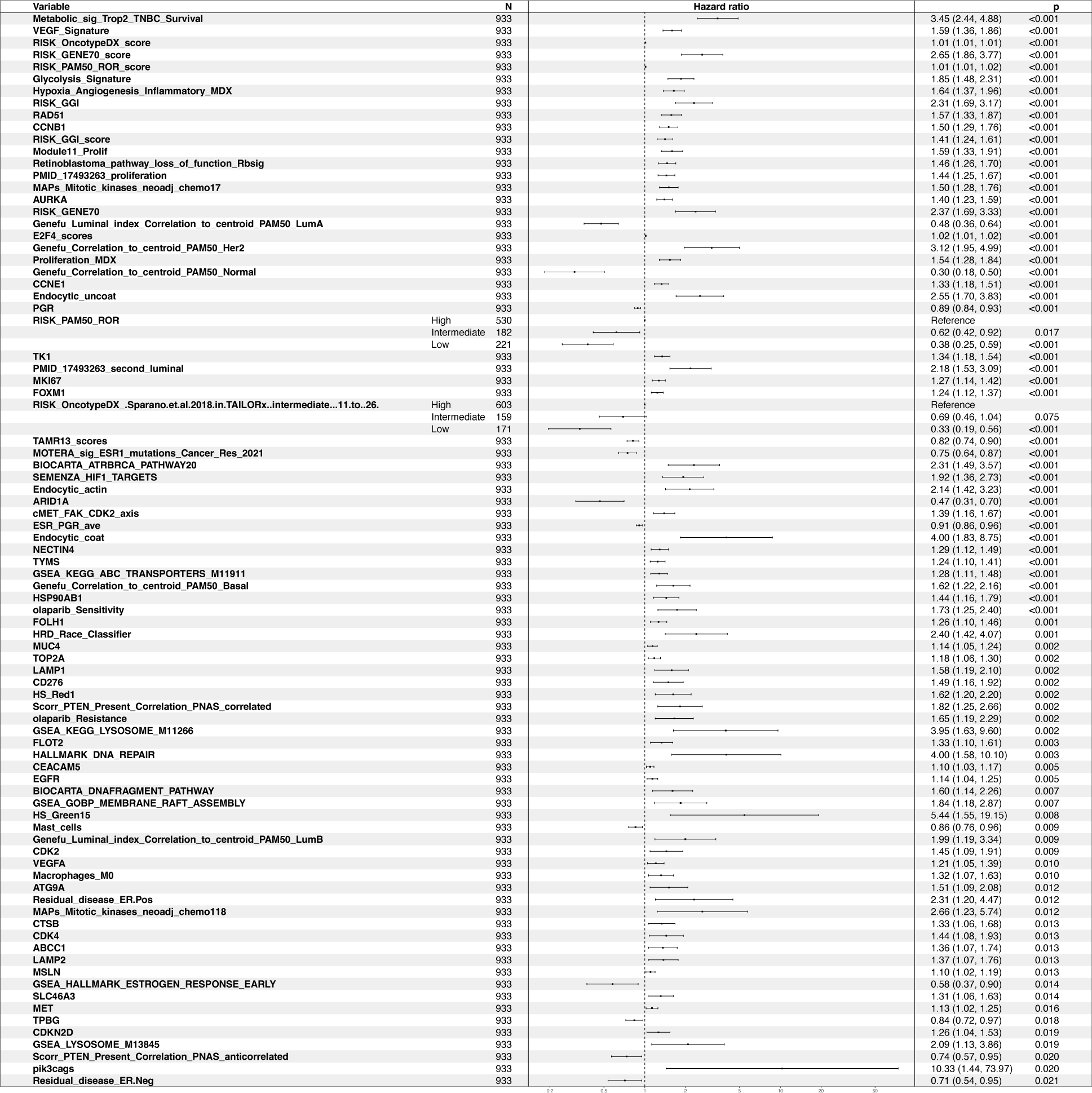
Genes and gene signatures associated with overall survival in the entire cohort. Forest plot illustrating significant Hazard ratios (HR) obtained from univariate Cox Proportional Hazard (Cox-PH) analyses, using a continuous signature scoring method calculated for the whole dataset (n=1015). Actual numbers in each analysis are lower due to missing follow up data and are denoted in the figure.

**Extended Figure 3.**
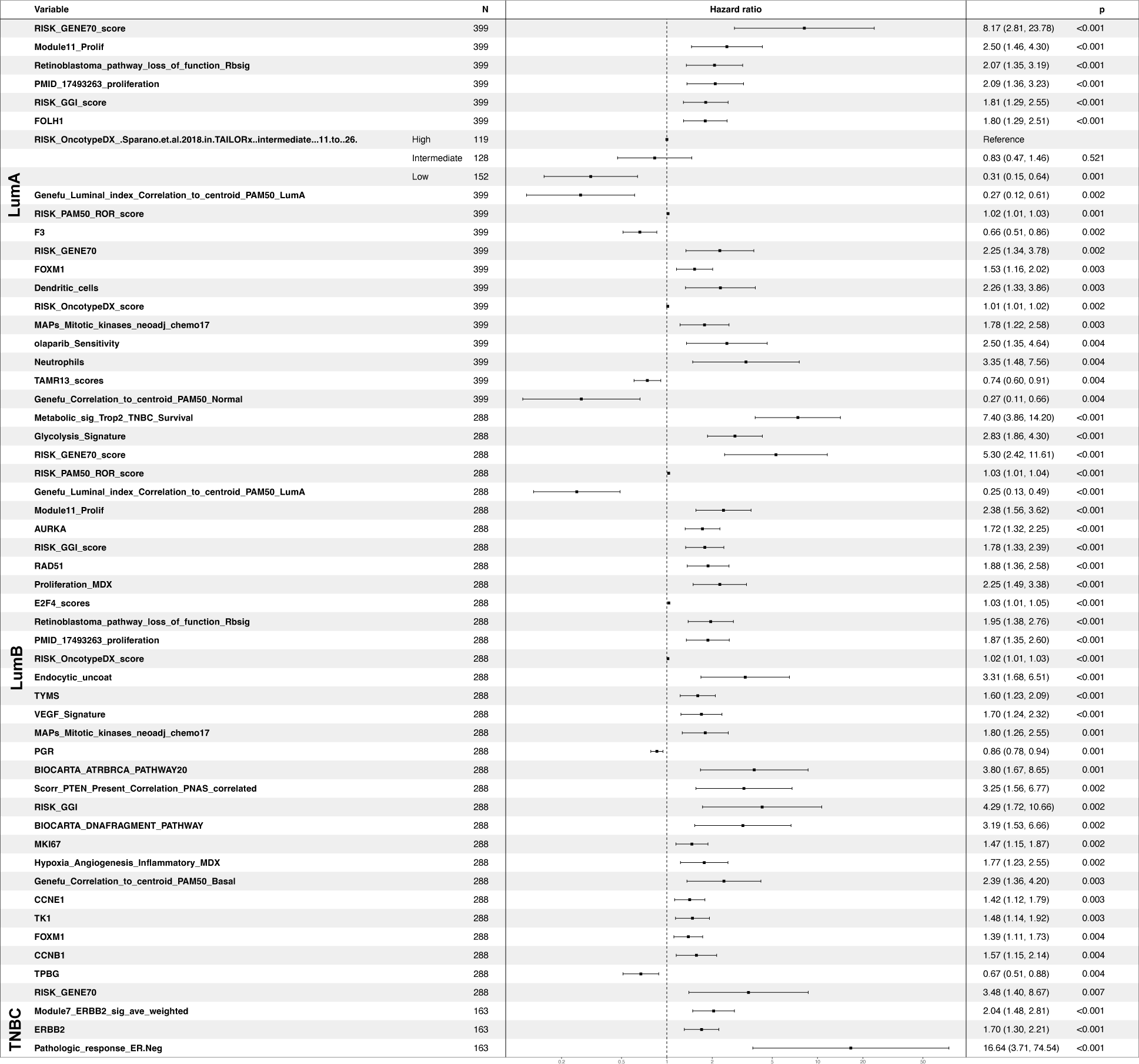
Gene signatures associated with overall survival in specific clinical subtypes. A Forest plot illustrating the significant Hazard ratios (HR) obtained from univariate Cox Proportional Hazard (Cox-PH) analyses, using categorical gene signature scoring method separately calculated for each biobank clinical subtype (n=1015). Actual numbers in each analysis are lower due to missing follow up data and are denoted in the figure.

**Extended Figure 4.**
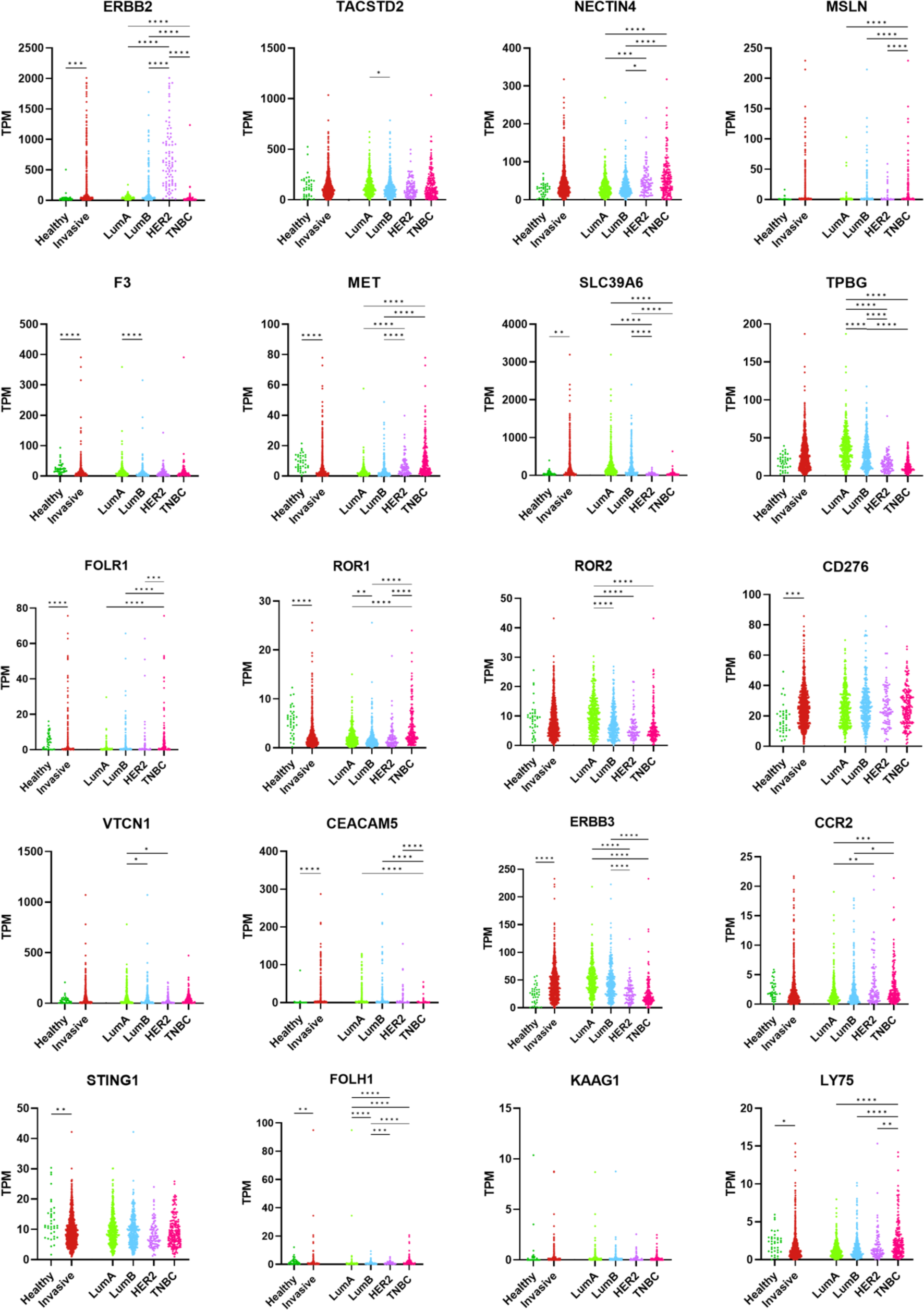
Expression of ADC targets in healthy versus invasive breast cancer tissue. Scatter dot plots illustrate the expression of ADC antigen targets assessed by RNA sequencing in healthy tissue, invasive breast cancer tissue, and individual clinical subtypes. *p < 0.05, **p < 0.01 ***p < 0.001, ****p < 0.0001.

**Extended Table 1.** Cohort descriptive statistics.

| Characteristic | Luminal A |  |  | Luminal B |  |  | Her2 |  |  | TNBC |  |  |
| --- | --- | --- | --- | --- | --- | --- | --- | --- | --- | --- | --- | --- |
| Subtype sample # | 432 |  |  | 313 |  |  | 87 |  |  | 181 |  |  |
| Biobank | PATH | GRAZ | Mal + AMS | PATH | GRAZ | Mal + AMS | PATH | GRAZ | Mal + AMS | PATH | GRAZ | Mal + AMS |
| Sample # | 205<br>(47%) | 205<br>(47%) | 22<br>(5%) | 185<br>(59%) | 125<br>(40%) | 3<br>(1%) | 61<br>(70%) | 23<br>(26%) | 3<br>(3%) | 141<br>(78%) | 37<br>(20%) | 3<br>(2%) |
| Sex |  |  |  |  |  |  |  |  |  |  |  |  |
| Female | 205<br>(100%) | 202<br>(99%) | - | 185<br>(100%) | 121<br>(97%) | - | 61<br>(100%) | 23<br>(100%) | - | 141<br>(100%) | 37<br>(100%) | - |
| Male | 0<br>(0%) | 3<br>(1%) | - | 0<br>(0%) | 4<br>(3%) | - | 0<br>(0%) | 0<br>(0%) | - | 0<br>(0%) | 0<br>(0%) | - |
| Median age at surgery (years) | 62 | 57 | 62 | 62 | 59 | 62 | 62 | 52 | 62 | 57 | 52 | 52 |
| Menopausal status |  |  |  |  |  |  |  |  |  |  |  |  |
| Premenopausal | 35<br>(17%) | - | - | 42<br>(23%) | - | - | 7<br>(11%) | - | - | 29<br>(21%) | - | - |
| Perimenopausal | 6<br>(3%) | - | - | 3<br>(2%) | - | - | 1<br>(2%) | - | - | 8<br>(6%) | - | - |
| Postmenopausal | 159<br>(78%) | - | - | 137<br>(74%) | - | - | 43<br>(70%) | - | - | 93<br>(66%) | - | - |
| Unavailable | 5<br>(2%) | - | - | 3<br>(2%) | - | - | 10<br>(16%) | - | - | 11<br>(8%) | - | - |
| Tumor size (T) |  |  |  |  |  |  |  |  |  |  |  |  |
| pT1 | 128<br>(62%) | 150<br>(73%) | 7<br>(32%) | 79<br>(43%) | 71<br>(56%) | 1<br>(33%) | 26<br>(43%) | 14<br>(61%) | 1<br>(33%) | 68<br>(48%) | 22<br>(59%) | 0<br>(0%) |
| pT2 | 69<br>(34%) | 45<sup>(22%)</sup> | 11<br>(50%) | 94<br>(51%) | 48<br>(38%) | 1<br>(33%) | 31<br>(51%) | 8<br>(35%) | 1<br>(33%) | 65<br>(46%) | 14<br>(38%) | 2<br>(67%) |
| pT3 + pT4 | 8<br>(4%) | 10<br>(5%) | 4<br>(18%) | 12<br>(6%) | 6<br>(5%) | 1<br>(33%) | 4<br>(7%) | 1<br>(4%) | 1<br>(33%) | 8<br>(6%) | 1<br>(3%) | 1<br>(33%) |
| Node status (N) |  |  |  |  |  |  |  |  |  |  |  |  |
| pN0 | 98<br>(48%) | 127<br>(62%) | 5<br>(23%) | 93<br>(50%) | 70<br>(56%) | 0<br>(0%) | 30<br>(49%) | 10<br>(43%) | 0<br>(0%) | 99<br>(70%) | 28<br>(76%) | 0<br>(0%) |
| pN1 | 88<br>(43%) | 57<br>(28%) | 11<br>(50%) | 65<br>(35%) | 36<br>(29%) | 1<br>(33%) | 21<br>(34%) | 6<br>(26%) | 0<br>(0%) | 29<br>(21%) | 8<br>(22%) | 0<br>(0%) |
| pN2 | 13<br>(6%) | 10<br>(5%) | 4<br>(18%) | 18<br>(10%) | 10<br>(8%) | 0<br>(0%) | 4<br>(7%) | 3<br>(13%) | 2<br>(67%) | 6<br>(4%) | 0<br>(0%) | 1<br>(33%) |
| pN3 | 6<br>(3%) | 8<br>(4%) | 1<br>(5%) | 9<br>(5%) | 8<br>(6%) | 1<br>(33%) | 5<br>(8%) | 2<br>(9%) | 0<br>(0%) | 7<br>(5%) | 1<br>(3%) | 1<br>(33%) |
| Unavailable | 0<br>(0%) | 3<br>(1%) | 1<br>(5%) | 0<br>(0%) | 1<br>(1%) | 1<br>(33%) | 1<br>(2%) | 2<br>(9%) | 1<br>(33%) | 0<br>(0%) | 0<br>(0%) | 1<br>(33%) |
| Grade (G) |  |  |  |  |  |  |  |  |  |  |  |  |
| G1 | 61<br>(30%) | 60<br>(29%) | 3<br>(14%) | 1<br>(1%) | 4<br>(3%) | 0<br>(0%) | 0<br>(0%) | 0<br>(0%) | 0<br>(0%) | 1<br>(1%) | 1<br>(3%) | 0<br>(0%) |
| G2 | 144<br>(70%) | 140<sup>1</sup><br>(68%) | 10<br>(45%) | 30<br>(16%) | 31<sup>2</sup><br>(25%) | 3<br>(100%) | 17<br>(28%) | 3<br>(13%) | 1<br>(33%) | 32<br>(23%) | 7<br>(19%) | 1<br>(33%) |
| G3 | 0<br>(0%) | 4<br>(2%) | 9<br>(41%) | 154<br>(83%) | 89<br>(71%) | 0<br>(0%) | 44<br>(72%) | 20<br>(87%) | 2<br>(67%) | 108<br>(77%) | 28<sup>3</sup><br>(76%) | 2<br>(67%) |

<sup>1</sup> One specimen classified as G2&G3
<sup>2</sup> Two specimens classified as G2&G3
<sup>3</sup> Two specimens classified as G4

|  |  |  |  |  |  |  |  |  |  |  |  |  |
| --- | --- | --- | --- | --- | --- | --- | --- | --- | --- | --- | --- | --- |
| Unavailable | 0<br>(0%) | 1<br>(<1%) | 0<br>(0%) | 0<br>(0%) | 1<br>(1%) | 0<br>(0%) | 0<br>(0%) | 0<br>(0%) | 0<br>(0%) | 0<br>(0%) | 1<br>(3%) | 0<br>(0%) |
| **Lymphatic invasion (L)** |  |  |  |  |  |  |  |  |  |  |  |  |
| L0 | 137<br>(67%) | - | 15<br>(68%) | 105<br>(57%) | - | 2<br>(67%) | 31<br>(51%) | - | 2<br>(67%) | 99<br>(70%) | - | 0<br>(0%) |
| L1 | 38<br>(19%) | - | 6<br>(27%) | 55<br>(30%) | - | 0<br>(0%) | 24<br>(39%) | - | 0<br>(0%) | 35<br>(25%) | - | 2<br>(67%) |
| Unavailable | 30<br>(15%) | - | 1<br>(5%) | 25<br>(14%) | - | 1<br>(33%) | 6<br>(10%) | - | 1<br>(33%) | 7<br>(5%) | - | 1<br>(33%) |
| **Residual tumor (PATH only)** |  |  |  |  |  |  |  |  |  |  |  |  |
| R0 | 204<br>(100%) | - | 19<br>(86%) | 180<br>(97%) | - | 2<br>(67%) | 60<br>(98%) | - | 2<br>(67%) | 138<br>(98%) | - | 2<br>(67%) |
| R1 | 0<br>(0%) | - | 2<br>(9%) | 4<br>(2%) | - | 0<br>(0%) | 1<br>(2%) | - | 0<br>(0%) | 2<br>(1%) | - | 0<br>(0%) |
| Unavailable | 1<br>(<1%) | - | 1<br>(5%) | 1<br>(1%) | - | 1<br>(33%) | 0<br>(0%) | - | 1<br>(33%) | 1<br>(1%) | - | 1<br>(33%) |
| **Histopathological tumor types** |  |  |  |  |  |  |  |  |  |  |  |  |
| Invasive ductal carcinoma (IDC) | 142<br>(69%) | 135<br>(66%) | 17<br>(77%) | 154<br>(83%) | 103<br>(82%) | 3<br>(100%) | 52<br>(85%) | 21<br>(91%) | 3<br>(100%) | 125<br>(89%) | 26<br>(70%) | 3<br>(100%) |
| IDC mixed | 3<br>(1%) | 10<br>(5%) | 2<br>(9%) | 10<br>(5%) | 3<br>(2%) | 0<br>(0%) | 6<br>(10%) | 0<br>(0%) | 0<br>(0%) | 7<br>(5%) | 2<br>(5%) | 0<br>(0%) |
| Invasive lobular carcinoma (ILC) | 38<br>(19%) | 42<br>(20%) | 3<br>(14%) | 16<br>(9%) | 14<br>(11%) | 0<br>(0%) | 1<br>(2%) | 1<br>(4%) | 0<br>(0%) | 1<br>(1%) | 1<br>(3%) | 0<br>(0%) |
| ILC mixed | 4<br>(2%) | 0<br>(0%) | 0<br>(0%) | 0<br>(0%) | 0<br>(0%) | 0<br>(0%) | 0<br>(0%) | 0<br>(0%) | 0<br>(0%) | 0<br>(0%) | 0<br>(0%) | 0<br>(0%) |
| Metaplastic carcinoma | 0<br>(0%) | 0<br>(0%) | 0<br>(0%) | 0<br>(0%) | 0<br>(0%) | 0<br>(0%) | 0<br>(0%) | 0<br>(0%) | 0<br>(0%) | 3<br>(2%) | 5<br>(14%) | 0<br>(0%) |
| Mucinous carcinoma | 9<br>(4%) | 8<br>(4%) | 0<br>(0%) | 1<br>(1%) | 2<br>(2%) | 0<br>(0%) | 0<br>(0%) | 0<br>(0%) | 0<br>(0%) | 0<br>(0%) | 0<br>(0%) | 0<br>(0%) |
| Other rare histopathological subtypes | 8<br>(4%) | 9<br>(4%) | 0<br>(0%) | 4<br>(2%) | 3<br>(2%) | 0<br>(0%) | 2<br>(3%) | 0<br>(0%) | 0<br>(0%) | 5<br>(4%) | 2<br>(5%) | 0<br>(0%) |
| Unavailable | 1<br>(<1%) | 1<br>(<1%) | 0<br>(0%) | 0<br>(0%) | 0<br>(0%) | 0<br>(0%) | 0<br>(0%) | 1<br>(4%) | 0<br>(0%) | 0<br>(0%) | 1<br>(3%) | 0<br>(0%) |
| **Estrogen receptor positive (%)** | 205<br>(100%) | - | - | 185<br>(100%) | - | - | 0<br>(0%) | - | - | 0<br>(0%) | - | - |
| IRS 0 | 0<br>(0%) | - | - | 0<br>(0%) | - | - | 41<br>(67%) | - | - | 97<br>(69%) | - | - |
| IRS 1-2 | 1<br>(<1%) | - | - | 23<br>(12%) | - | - | 2<br>(3%) | - | - | 4<br>(3%) | - | - |
| IRS 3-4 | 4<br>(2%) | - | - | 20<br>(11%) | - | - | 0<br>(0%) | - | - | 0<br>(0%) | - | - |
| IRS 6-8 | 27<br>(13%) | - | - | 25<br>(14%) | - | - | 0<br>(0%) | - | - | 0<br>(0%) | - | - |
| IRS 9 | 18<br>(9%) | - | - | 20<br>(11%) | - | - | 0<br>(0%) | - | - | 0<br>(0%) | - | - |
| IRS 12 | 155<br>(76%) | - | - | 95<br>(51%) | - | - | 0<br>(0%) | - | - | 0<br>(0%) | - | - |
| Unavailable | 0<br>(0%) | - | - | 2<br>(1%) | - | - | 18<br>(30%) | - | - | 40<br>(28%) | - | - |
| **Progesterone receptor positive (%)** | 181<br>(88%) | - | - | 130<br>(70%) | - | - | 0<br>(0%) | - | - | 0<br>(0%) | - | - |
| IRS 0 | 20<br>(10%) | - | - | 43<br>(23%) | - | - | 43<br>(70%) | - | - | 97<br>(69%) | - | - |
| IRS 1-2 | 12<br>(6%) | - | - | 29<br>(16%) | - | - | 0<br>(0%) | - | - | 5<br>(4%) | - | - |
| IRS 3-4 | 24<br>(12%) | - | - | 23<br>(12%) | - | - | 0<br>(0%) | - | - | 0<br>(0%) | - | - |
| IRS 6-8 | 37<br>(18%) | - | - | 34<br>(18%) | - | - | 0<br>(0%) | - | - | 0<br>(0%) | - | - |
| IRS 9 | 34<br>(17%) | - | - | 21<br>(11%) | - | - | 0<br>(0%) | - | - | 0<br>(0%) | - | - |
| IRS 12 | 78<br>(38%) | - | - | 33<br>(18%) | - | - | 0<br>(0%) | - | - | 0<br>(0%) | - | - |
| Unavailable | 0<br>(0%) | - | - | 2<br>(1%) | - | - | 18<br>(30%) | - | - | 39<br>(28%) | - | - |
| **HER2 positive** | 0<br>(0%) | - | - | 51<br>(28%) | - | - | 61<br>(100%) | - | - | 0<br>(0%) | - | - |
| HER2 0 | 104<br>(51%) | - | - | 65<br>(35%) | - | - | 0<br>(0%) | - | - | 105<br>(74%) | - | - |
| HER2 1+ | 90<br>(44%) | - | - | 58<br>(31%) | - | - | 0<br>(0%) | - | - | 32<br>(23%) | - | - |
| HER2 2+ Neg. | 10<br>(5%) | - | - | 8<br>(4%) | - | - | 0<br>(0%) | - | - | 1<br>(1%) | - | - |
| HER2 2+ Pos. | 0<br>(0%) | - | - | 8<br>(4%) | - | - | 5<br>(8%) | - | - | 0<br>(0%) | - | - |
| HER2 3+ | 0<br>(0%) | - | - | 40<br>(22%) | - | - | 53<br>(87%) | - | - | 0<br>(0%) | - | - |
| Unavailable | 1<br>(<1%) | - | - | 6<br>(3%) | - | - | 3<br>(5%) | - | - | 3<br>(2%) | - | - |
| **KI67 positive** |  |  |  |  |  |  |  |  |  |  |  |  |
| ≤5% | 13<br>(6%) | 46<br>(22%) | - | 1<br>(1%) | 3<br>(2%) | - | 0<br>(0%) | 0<br>(0%) | - | 1<br>(1%) | 0<br>(0%) | - |
| >5 to <30% | 63<br>(31%) | 108<br>(52%) | - | 22<br>(12%) | 19<br>(15%) | - | 1<br>(2%) | 2<br>(9%) | - | 3<br>(2%) | 2<br>(5%) | - |
| ≥30% | 3<br>(1%) | 0<br>(0%) | - | 35<br>(19%) | 70<br>(56%) | - | 4<br>(7%) | 11<br>(48%) | - | 21<br>(15%) | 22<br>(59%) | - |
| Unavailable | 126<br>(61%) | 51<br>(25%) | - | 127<br>(69%) | 33<br>(26%) | - | 56<br>(92%) | 10<br>(43%) | - | 116<br>(82%) | 13<br>(35%) | - |
| **Treatment type** |  |  |  |  |  |  |  |  |  |  |  |  |
| **Chemotherapy** |  |  |  |  |  |  |  |  |  |  |  |  |
| Yes | 51<br>(25%) | 41<br>(20%) | - | 136<br>(74%) | 73<br>(58%) | - | 54<br>(89%) | 19<br>(83%) | - | 125<br>(89%) | 35<br>(95%) | - |
| No | 152<br>(74%) | 164<br>(80%) | - | 48<br>(26%) | 52<br>(41%) | - | 5<br>(8%) | 4<br>(17%) | - | 14<br>(10%) | 2<br>(5%) | - |
| Unavailable | 2<br>(1%) | 0<br>(0%) | - | 1<br>(1%) | 0<br>(0%) | - | 2<br>(3%) | 0<br>(0%) | - | 2<br>(1%) | 0<br>(0%) | - |
| **Anti-hormonal therapy** |  |  |  |  |  |  |  |  |  |  |  |  |
| Yes | 194<br>(95%) | 100<br>(49%) | - | 163<br>(88%) | 61<br>(48%) | - | 10<br>(16%) | 0<br>(0%) | - | 9<br>(6%) | 0<br>(0%) | - |
| No | 9<br>(4%) | 106<br>(51%) | - | 21<br>(11%) | 65<br>(52%) | - | 49<br>(80%) | 23<br>(100%) | - | 131<br>(93%) | 37<br>(100%) | - |
| Unavailable | 2<br>(1%) | 0<br>(0%) | - | 1<br>(1%) | 0<br>(0%) | - | 2<br>(3%) | 0<br>(0%) | - | 1<br>(1%) | 0<br>(0%) | - |
| **AI inhibitor therapy** |  |  |  |  |  |  |  |  |  |  |  |  |
| Yes | 129<br>(63%) | 101<br>(49%) | - | 123<br>(66%) | 54<br>(43%) | - | 9<br>(15%) | 0<br>(0%) | - | 5<br>(4%) | 0<br>(0%) | - |
| No | 72<br>(35%) | 104<br>(51%) | - | 59<br>(32%) | 71<br>(57%) | - | 52<br>(85%) | 23<br>(100<br>%) | - | 136<br>(96%) | 37<br>(100<br>%) | - |
| Unavailable | 4<br>(2%) | 0<br>(0%) | - | 3<br>(2%) | 0<br>(0%) | - | 0<br>(0%) | 0<br>(0%) | - | 0<br>(0%) | 0<br>(0%) | - |
| Anti-Her2 therapy |  |  |  |  |  |  |  |  |  |  |  |  |
| Yes | 0<br>(0%) | 0<br>(0%) | - | 42<br>(23%) | 20<br>(16%) | - | 50<br>(82%) | 22<br>(96%) | - | 1<br>(1%) | 0<br>(0%) | - |
| No | 203<br>(99%) | 205<br>(100<br>%) | - | 140<br>(76%) | 105<br>(83%) | - | 9<br>(15%) | 1<br>(4%) | - | 139<br>(99%) | 37<br>(100<br>%) | - |
| Unavailable | 2<br>(1%) | 0<br>(0%) | - | 3<br>(2%) | 0<br>(0%) | - | 2<br>(3%) | 0<br>(0%) | - | 1<br>(1%) | 0<br>(0%) | - |
| Radiotherapy |  |  |  |  |  |  |  |  |  |  |  |  |
| IORT | 6<br>(3%) | 0<br>(0%) | - | 2<br>(1%) | 0<br>(0%) | - | 0<br>(0%) | 0<br>(0%) | - | 1<br>(1%) | 0<br>(0%) | - |
| N | 18<br>(9%) | 40<br>(19%) | - | 14<br>(8%) | 21<br>(17%) | - | 4<br>(7%) | 5<br>(22%) | - | 15<br>(11%) | 5<br>(14%) | - |
| Y | 147<br>(72%) | 165<br>(80%) | - | 131<br>(71%) | 104<br>(83%) | - | 51<br>(84%) | 18<br>(78%) | - | 114<br>(81%) | 32<br>(86%) | - |
| Y + IORT | 31<br>(15%) | 0<br>(0%) | - | 37<br>(20%) | 0<br>(0%) | - | 5<br>(8%) | 0<br>(0%) | - | 10<br>(7%) | 0<br>(0%) | - |
| Unavailable | 3<br>(1%) | 0<br>(0%) | - | 1<br>(1%) | 0<br>(0%) | - | 1<br>(2%) | 0<br>(0%) | - | 1<br>(1%) | 0<br>(0%) | - |
| Metastasis |  |  |  |  |  |  |  |  |  |  |  |  |
| Yes | 8<br>(4%) | 35<br>(17%) | - | 12<br>(6%) | 41<br>(33%) | - | 3<br>(5%) | 7<br>(30%) | - | 18 | 5 | - |
| No | 173<br>(84%) | 170<br>(83%) | - | 120<br>(65%) | 84<br>(67%) | - | 48<br>(79%) | 16<br>(70%) | - | 93 | 32 | - |
| Unavailable/Lost<br>to follow-up | 24<br>(12%) | 0<br>(0%) | - | 53<br>(29%) | 0<br>(0%) | - | 10<br>(16%) | 0<br>(0%) | - | 30 | 0<br>(0%) | - |
| Progression free<br>survival (Median) | 70<br>mont<br>hs | 90<br>mont<br>hs | - | 64.5<br>mont<br>hs | 94<br>mont<br>hs | - | 98.5<br>mont<br>hs | 89<br>mont<br>hs | - | 84.5<br>mont<br>hs | 123<br>mont<br>hs | - |
| Unavailable/Lost<br>to follow-up | 8<br>(4%) | 1<br>(<1%) | - | 23<br>(12%) | 2<br>(2%) | - | 3<br>(5%) | 0<br>(0%) | - | 19 | 0<br>(0%) | - |
| Overall survival<br>(Median) | 72<br>mont<br>hs | 97<br>mont<br>hs | - | 66<br>mont<br>hs | 103<br>mont<br>hs | - | 102<br>mont<br>hs | 101<br>mont<br>hs | - | 85.5<br>mont<br>hs | 123<br>mont<br>hs | - |
| Unavailable/Lost<br>to follow-up | 11<br>(5%) | 0<br>(0%) | - | 22<br>(12%) | 0<br>(0%) | - | 1<br>(2%) | 0<br>(0%) | - | 15 | 0<br>(0%) | - |
| Deceased |  |  |  |  |  |  |  |  |  |  |  |  |
| Yes | 40<br>(20%) | 25<br>(12%) | - | 61<br>(33%) | 19<br>(15%) | - | 17<br>(28%) | 6<br>(26%) | - | 50<br>(35%) | 3<br>(8%) | - |
| No | 165<br>(80%) | 0<br>(0%) | - | 124<br>(67%) | 0<br>(0%) | - | 44<br>(72%) | 0<br>(0%) | - | 91<br>(65%) | 0<br>(0%) | - |
| Unavailable | 0<br>(0%) | 180<br>(88%) | - | 0<br>(0%) | 106<br>(85%) | - | 0<br>(0%) | 17<br>(74%) | - | 0<br>(0%) | 34<br>(92%) | - |

**Extended Table 2.** Overview of differential gene expression analysis among molecular subtypes.

| Groups | DESeq2 (LFC shrinkage) | Wilcoxon rank-sum test | Intersection |
| --- | --- | --- | --- |
| Her2onl vs TNBC<br>*Downregulated*<br>*Upregulated* | 1138<br>*590*<br>*548* | 1372<br>*704*<br>*668* | 1006<br>*523*<br>*483* |
| Her2onl vs LumA<br>*Downregulated*<br>*Upregulated* | 3636<br>*1188*<br>*2448* | 4784<br>*1312*<br>*3472* | 3364<br>*1094*<br>*2270* |
| Her2onl vs LumB<br>*Downregulated*<br>*Upregulated* | 1577<br>*621*<br>*956* | 2181<br>*734*<br>*1447* | 1338<br>*547*<br>*791* |
| LumA vs TNBC<br>*Downregulated*<br>*Upregulated* | 4950<br>*3330*<br>*1620* | 5568<br>*3902*<br>*1666* | 4661<br>*3149*<br>*1512* |
| LumB vs TNBC<br>*Downregulated*<br>*Upregulated* | 2635<br>*1564*<br>*1071* | 2954<br>*1785*<br>*1169* | 2356<br>*1360*<br>*996* |
| LumA vs LumB<br>*Downregulated*<br>*Upregulated* | 746<br>*621*<br>*125* | 802<br>*686*<br>*116* | 525<br>*443*<br>*82* |

**Extended Table 3.** Overview of differential gene expression analysis among *ERBB2* positive, low, and negative samples stratified by *ESR1* status.

|  | Groups | DESeq2 (LFC shrinkage) | Wilcoxon rank-sum test | Intersection |
| --- | --- | --- | --- | --- |
| ERneg | Her2low vs HER2neg<br>*Downregulated*<br>*Upregulated* | 851<br>*785*<br>*66* | 682<br>*601*<br>*81* | 330<br>*318*<br>*12* |
|  | Her2low vs HER2pos<br>*Downregulated*<br>*Upregulated* | 1051<br>*240*<br>*811* | 1999<br>*868*<br>*1131* | 727<br>*207*<br>*520* |
|  | Her2neg vs HER2pos<br>*Downregulated*<br>*Upregulated* | 2427<br>*412*<br>*2015* | 2310<br>*923*<br>*1387* | 1073<br>*355*<br>*718* |
| ERpos | Her2low vs HER2neg<br>*Downregulated*<br>*Upregulated* | 5436<br>*5428*<br>*8* | 4779<br>*4759*<br>*20* | 3180<br>*3178*<br>*2* |
|  | Her2low vs HER2pos<br>*Downregulated*<br>*Upregulated* | 131<br>*62*<br>*69* | 141<br>*85*<br>*56* | 37<br>*29*<br>*8* |
|  | Her2neg vs HER2pos<br>*Downregulated*<br>*Upregulated* | 7497<br>*84*<br>*7413* | 603<br>*76*<br>*527* | 396<br>*48*<br>*348* |

