## Supplementary information for "Multiplexed RNA-FISH-guided Laser Capture Microdissection RNA Sequencing Improves Breast Cancer Molecular Subtyping, Prognostic Classification, and Predicts Response to Antibody Drug Conjugates"

**Short title:** Multiplexed RNA-FISH-guided LCM-seq

**Authors:**

Evan D. Paul[1,2], Barbora Huraiová[1,2], Natália Valková[1,2,3], Natalia Birknerova[1,2], Daniela Gábrišová[1,2], Sona Gubova[1,2], Helena Ignačáková[1,2], Tomáš Ondris[1,2], Silvia Bendíková[1,2], Jarmila Bíla[1,2.], Katarína Buranovská[1,2], Diana Drobná[1,2], Zuzana Krchnakova[1,2], Maryna Kryvokhyzha[1,2], Daniel Lovíšek[1,2], Viktoriia Mamoilyk[1,2], Veronika Mančíková[1,2], Nina Vojtaššáková[1,2], Michaela Ristová[1,2,4], Iñaki Comino-Méndez[5], Igor Andrašina[6], Pavel Morozov[7], Thomas Tuschl[7], Fresia Pareja[8,*], Pavol Čekan[1,2,*]

### Supplementary information

#### Supplementary Figure 1.

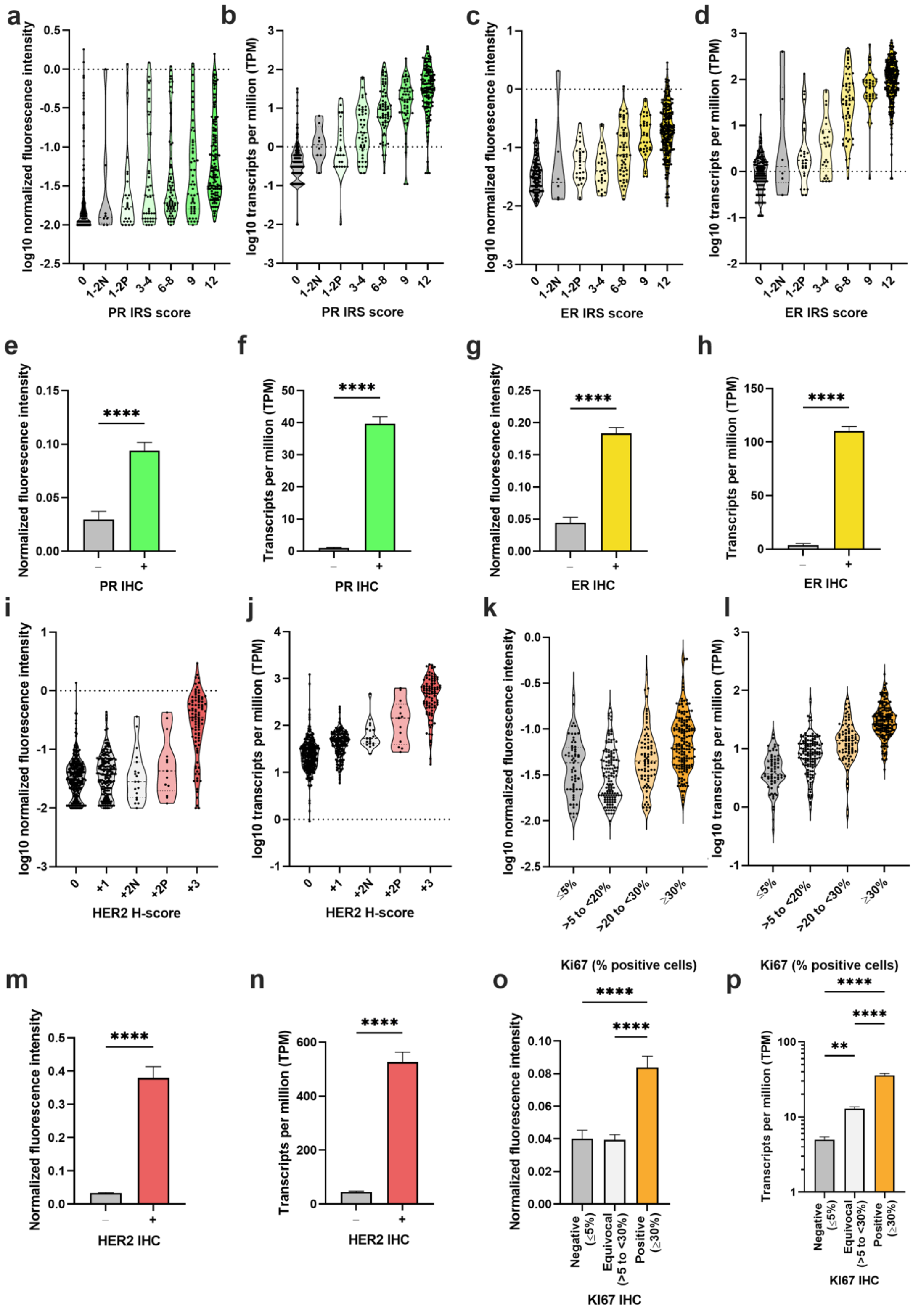

**Supplementary Figure 1. Comparison of RNA-FISH/-SEQ with biobank IHC results.** Graphs show the comparison of RNA-FISH and RNA-SEQ to biobank IHC results for PR (**a,b,e,f**), ER (**c,d,g,f**), HER2 (**i,j,m,n**), and Ki67 (**k,l,o,p**). Data are presented as a comparison to either IRS score for PR/ER, H-Score for HER2, percent of positive cells for Ki67, or as negative or positive IHC expression. Dotted lines in the violin plots show the median and quartiles. $^{*}p < 0.05$, $^{**}p < 0.01$ $^{***}p < 0.001$, $^{****}p < 0.0001$.

**Supplementary Figure 2.**

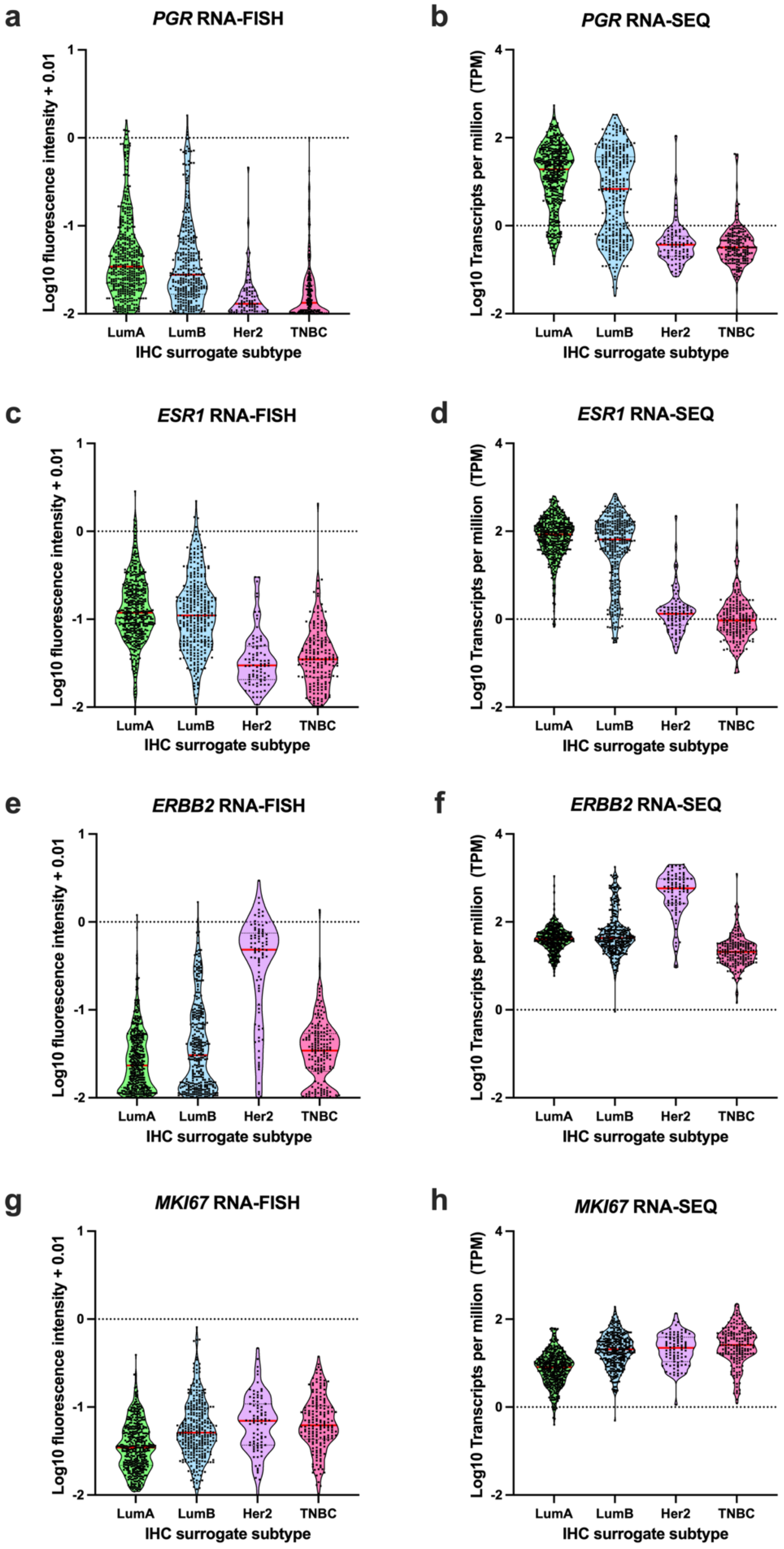

**Supplementary Figure 2. Expression of biomarkers as quantified by RNA-FISH and RNA-SEQ in IHC clinical breast cancer subtypes.** Violin plots show PR (**a,b**), ER (**c,d**), HER2 (**e,f**), and Ki67 (**g,h**) expression in each molecular subtype, as determined by IHC biobank results according to the 2013 St. Gallen classification of intrinsic subtypes, quantified by either RNA-FISH (**a,c,e,g**) or RNA-SEQ (**b,d,f,h**). Each dot represents an individual sample and the lines in each violin plot denote the median and quartiles (25% and 75%).

**Supplementary Figure 3.**

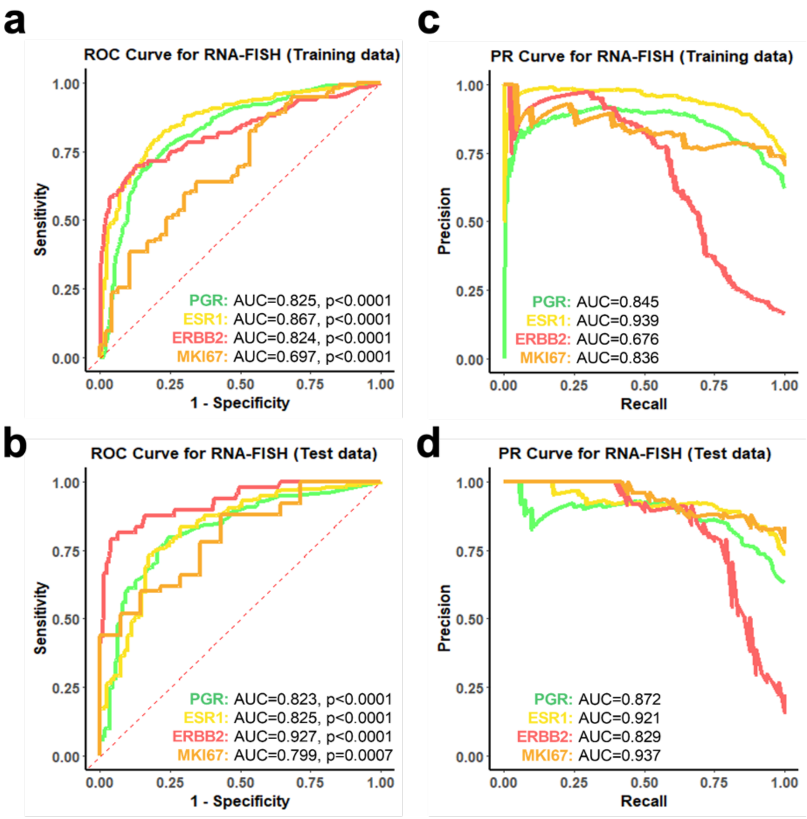


| SI thresholds (Maximum Cohens' K) | |
|---|---|
| PGR RNA-FISH | ≥ 0.011 |
| ESR1 RNA-FISH | ≥ 0.038 |
| ERBB22 RNA-FISH | ≥ 0.123 |
| MKI67 RNA-FISH | ≥ 0.05 |

**Supplementary Figure 3. Diagnostic performance of RNA-FISH compared to IHC.** ROC (**a**, **b**) and PR (**c**, **d**) curves from the 70:30 training (**a**, **c**) and test (**b**, **d**) datasets illustrate the performance of RNA-FISH at classifying biomarker expression for *PGR*, *ESR1*, *ERBB2*, and *MKI67*. The bottom table shows the RNA-FISH signal intensity (SI) thresholds for each marker derived from the training set by identifying the threshold value that maximized agreement (Cohen's K) between biobank IHC results. These threshold values were then prespecified for use in the test dataset. AUC, area under the curve; *ERBB2*, Erb-B2 Receptor Tyrosine Kinase 2 gene (HER2); *ESR1*, estrogen receptor 1 gene; *MKI67*, marker of proliferation Ki-67 gene; *PGR*, progesterone receptor gene. For Ki-67, ≤5% and ≥30% were used as cutoff values for negativity and positivity, respectively.

**Supplementary Figure 4.**

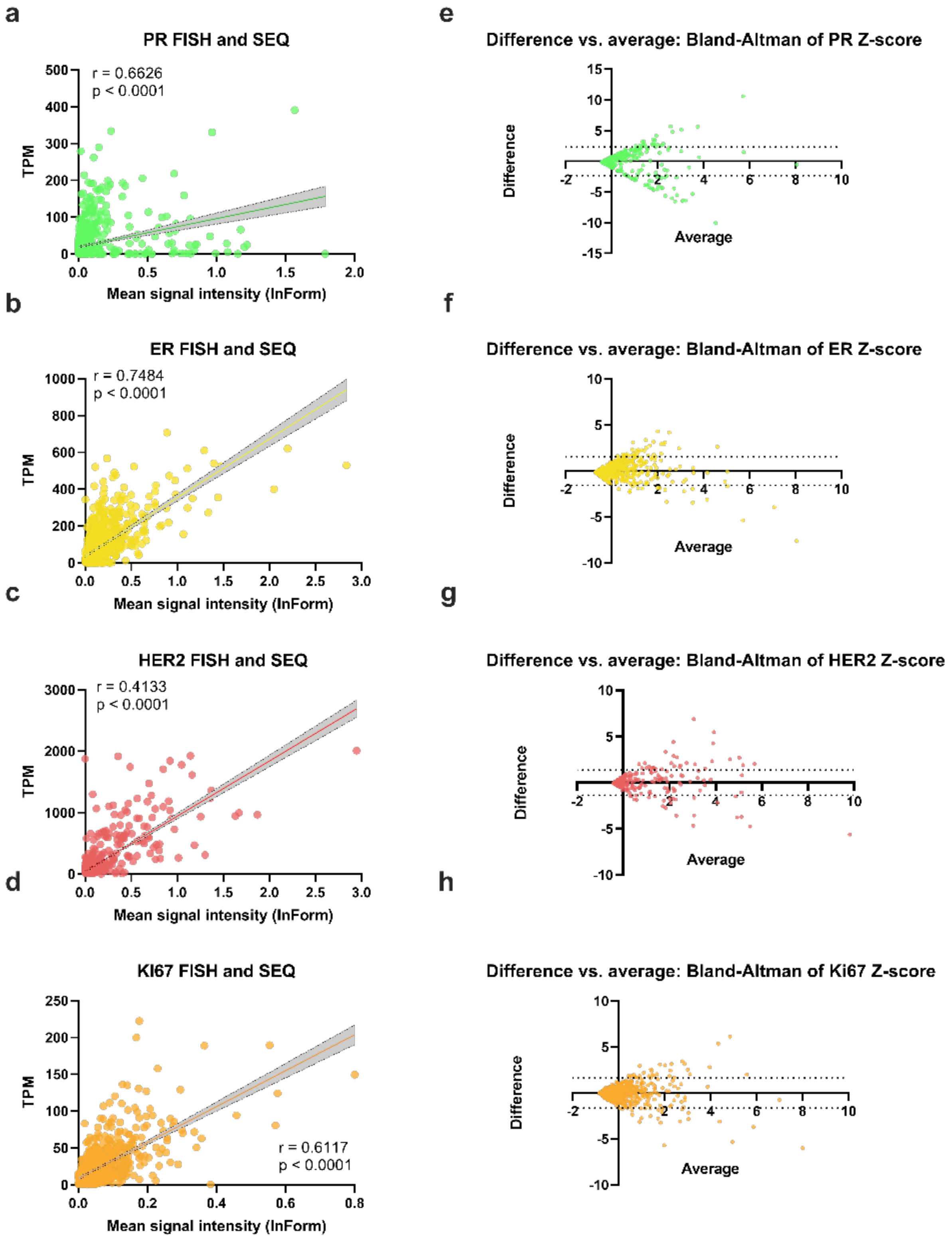

**Supplementary Figure 4. Concordance of biomarker expression between RNA-FISH and RNA-SEQ.** (**a-d**) Graphs depict correlations between RNA-FISH (mean signal intensity) and RNA-SEQ (transcripts per million, TPM) for PR (**a**), ER (**b**), HER2 (**c**), and KI67 (**d**). Correlations are Spearman's rho and the shaded region represents the 95% confidence bands of the best fit linear regression line. (**e-h**) Bland Altman plots (using difference vs average of normalized Z scores) illustrating agreement and potential biases of RNA-FISH and RNA-SEQ. Dotted lines show the 95% limits of agreement.

**Supplementary Figure 5.**

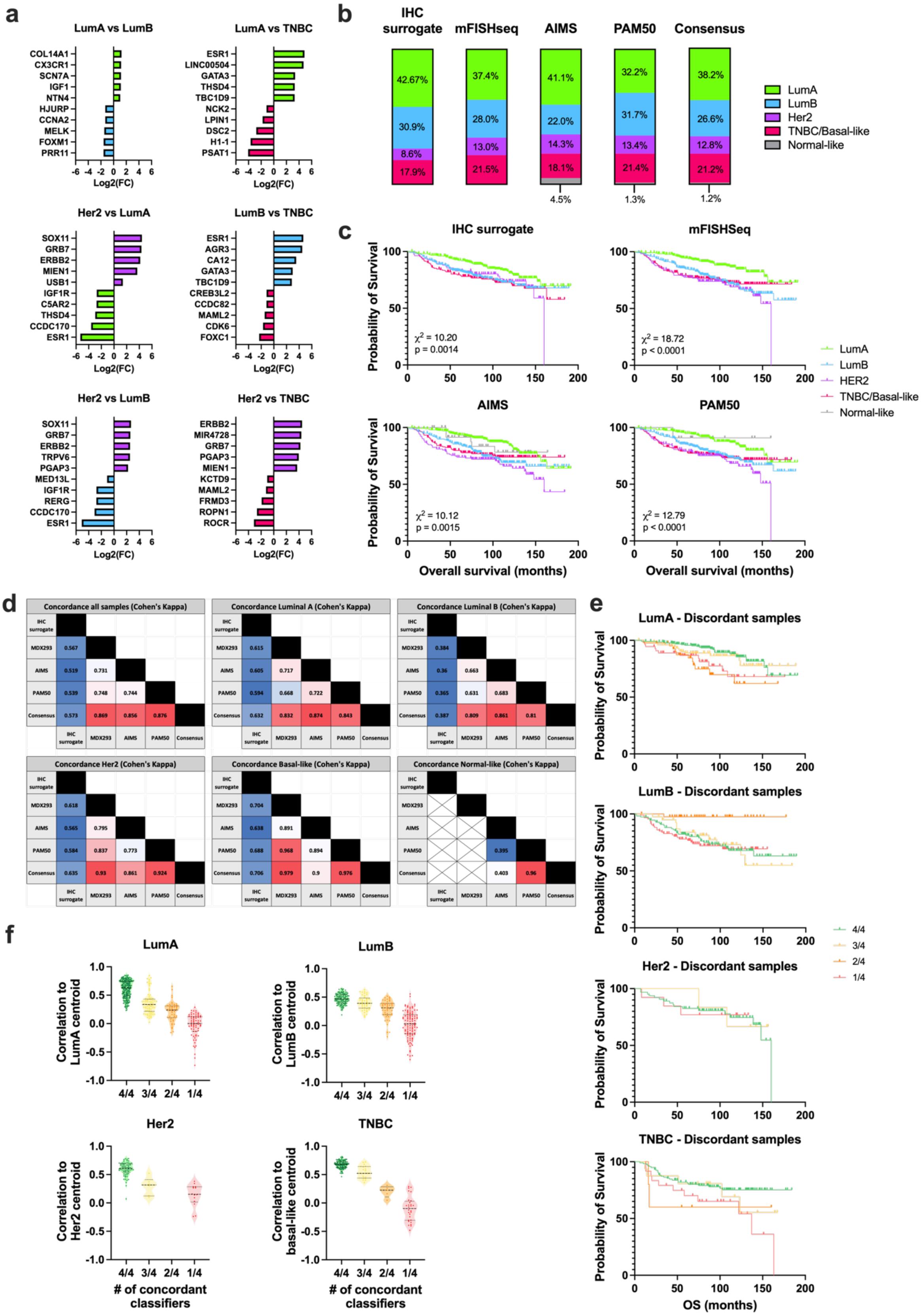

**Supplementary Figure 5. Comparison of molecular subtyping schemes in terms of survival and single sample concordance.** (**a**) Graphs show the top 5 differentially expressed genes (log2 fold change) that are either upregulated or downregulated between each molecular subtype. The data are arranged such that the direction of the log2 fold change relates to the first subtype listed in the graphs title (i.e., LumA vs LumB comparison shows genes that are upregulated and downregulated in Luminal A samples relative to Luminal B samples). (**b**) Proportion of samples classified by each classifier into the respective IHC surrogate or intrinsic molecular subtypes. (**c**) Kaplan-Meier survival curves comparing overall survival by molecular subtype according to the IHC surrogate, mFISHseq, AIMS, or PAM50 subtyping schemes. (**d**) Cohen's kappa concordance between IHC surrogate, mFISHseq, AIMS, PAM50, consensus subtyping schemes for all samples and samples stratified by subtype. (**e**) Survival curves illustrating probability of overall survival stratified by IHC surrogate subtype for samples that showed perfect concordance between all four classifiers (4/4, IHC surrogate, mFISHseq, AIMS, and PAM50) as well as discordant samples that showed concordance between three out of four classifiers (3/4, IHC surrogate and any two multigene classifiers), two out of four classifiers (2/4, IHC surrogate and any one multigene classifier), or only the IHC surrogate classifier (1/4). (**f**) Metrics of instability as assessed by correlation to the corresponding PAM50 subtype centroid for samples with perfect concordance (4/4) or samples showing discordance in one or more classifiers (3/3, 2/4, and 1/4) stratified by IHC surrogate subtype.

**Supplementary Figure 6.**

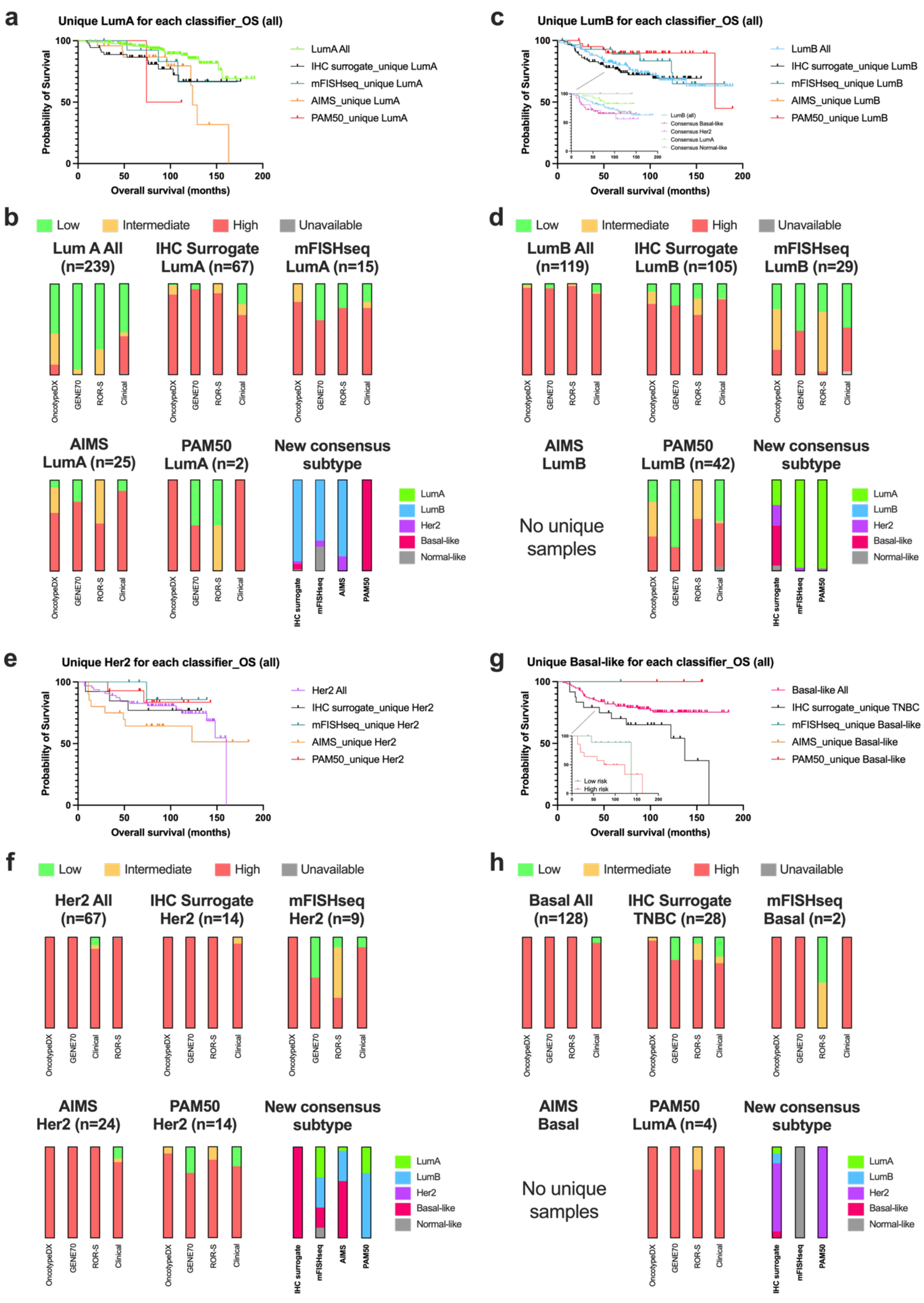

**Supplementary Figure 6. Single sample discordance and reclassification by consensus subtyping.** Overall survival of samples uniquely classified by only one subtyping approach (IHC surrogate, mFISHseq, AIMS, and PAM50) as luminal A (**a**), luminal B (**c**), Her2 (**e**), and TNBC/basal-like (**g**). The proportion of samples with low, intermediate, or high genomic risk assessed by OncotypeDX, GENE70, and ROR-S as well as clinical risk in samples uniquely classified as luminal A (**b**), luminal B (**d**), Her2 (**f**), and TNBC/basal-like (**h**). The uniquely classified samples that were reclassified to a new consensus molecular subtype are depicted in the bottom right bar charts of **b**, **d**, **f**, and **h**.

**Supplementary Figure 7.**

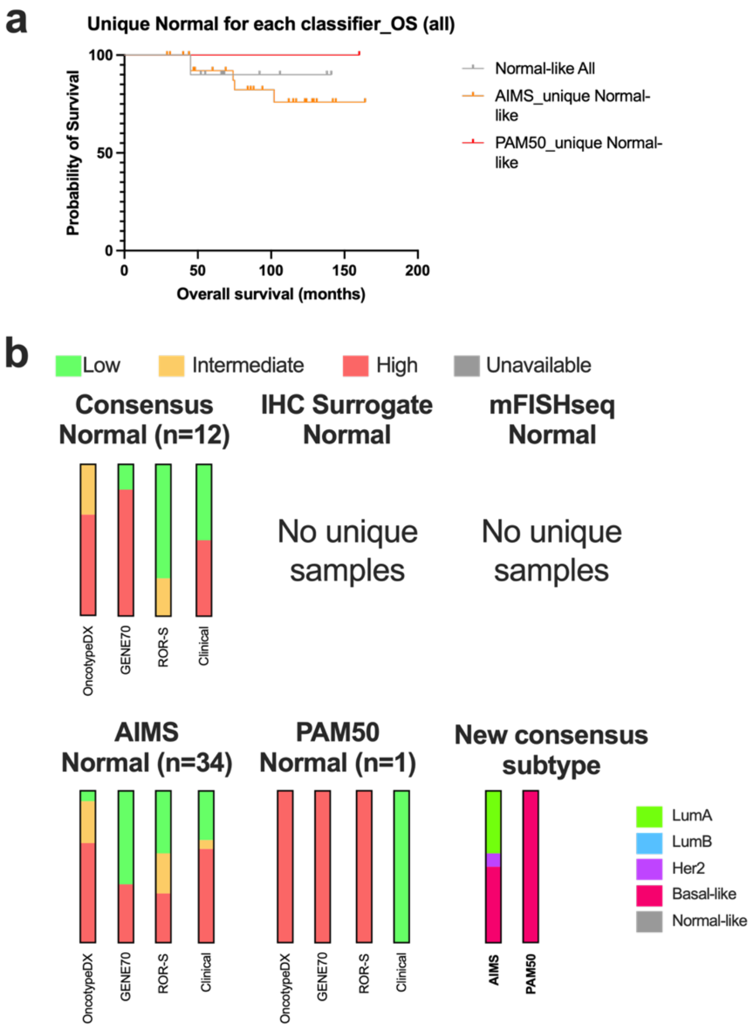


**Supplementary Figure 7. Single sample discordance for the normal-like subtype and reclassification by consensus subtyping.** (**a**) Overall survival of samples uniquely classified as normal-like by only one subtyping approach (AIMS, and PAM50). (**b**) The proportion of normal-like samples with low, intermediate, or high genomic risk assessed by OncotypeDX, GENE70, and ROR-S as well as clinical risk. (b, bottom right panel) The uniquely classified normal-like samples that were reclassified to a new consensus molecular subtype.

**Supplementary Figure 8.**

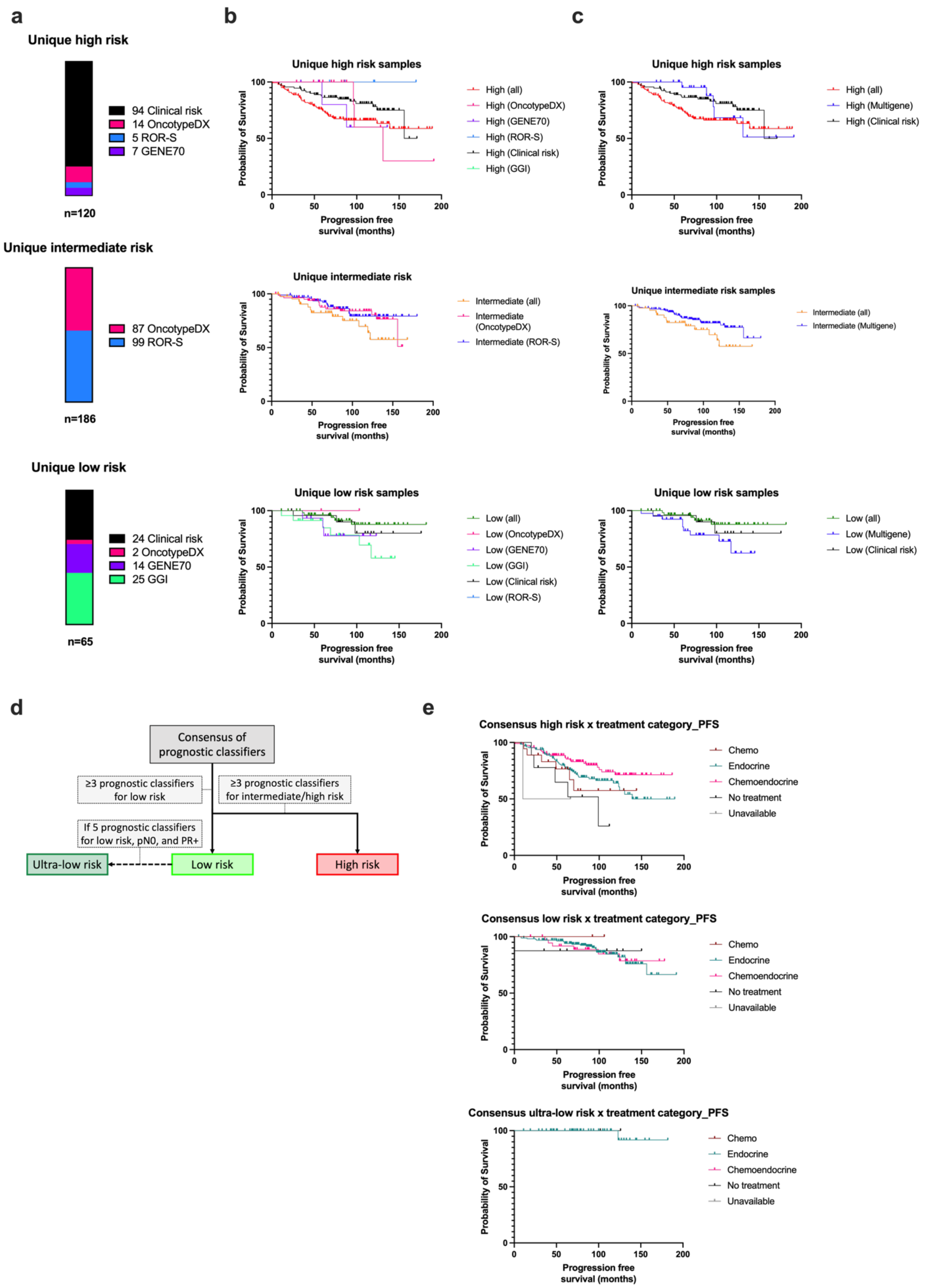

**Supplementary Figure 8: Development of consensus prognostic risk categories** (**a**) Proportion of uniquely classified patients (i.e., patients classified in a risk category by only one classifier) in high, intermediate, and low risk categories. (**b**) Kaplan-Meier (KM) progression free survival (PFS) curves showing patients uniquely classified by an individual classifier into high (top panel), intermediate (middle panel), and low (bottom panel) risk categories. (**c**) The same PFS curves in (**b**) after consolidating unique classifications from OncotypeDX, ROR-S, GENE70, and GGI into a single Multigene category for each risk category. (**d**) Exploratory decision tree describing criteria for consensus classification of patients into high, low, and ultra-low risk categories. (**e**) KM curves for high (top panel), low (middle panel), and ultra-low (bottom panel) consensus risk categories depicting PFS after stratifying patients into treatment categories.

**Supplementary Figure 9.**

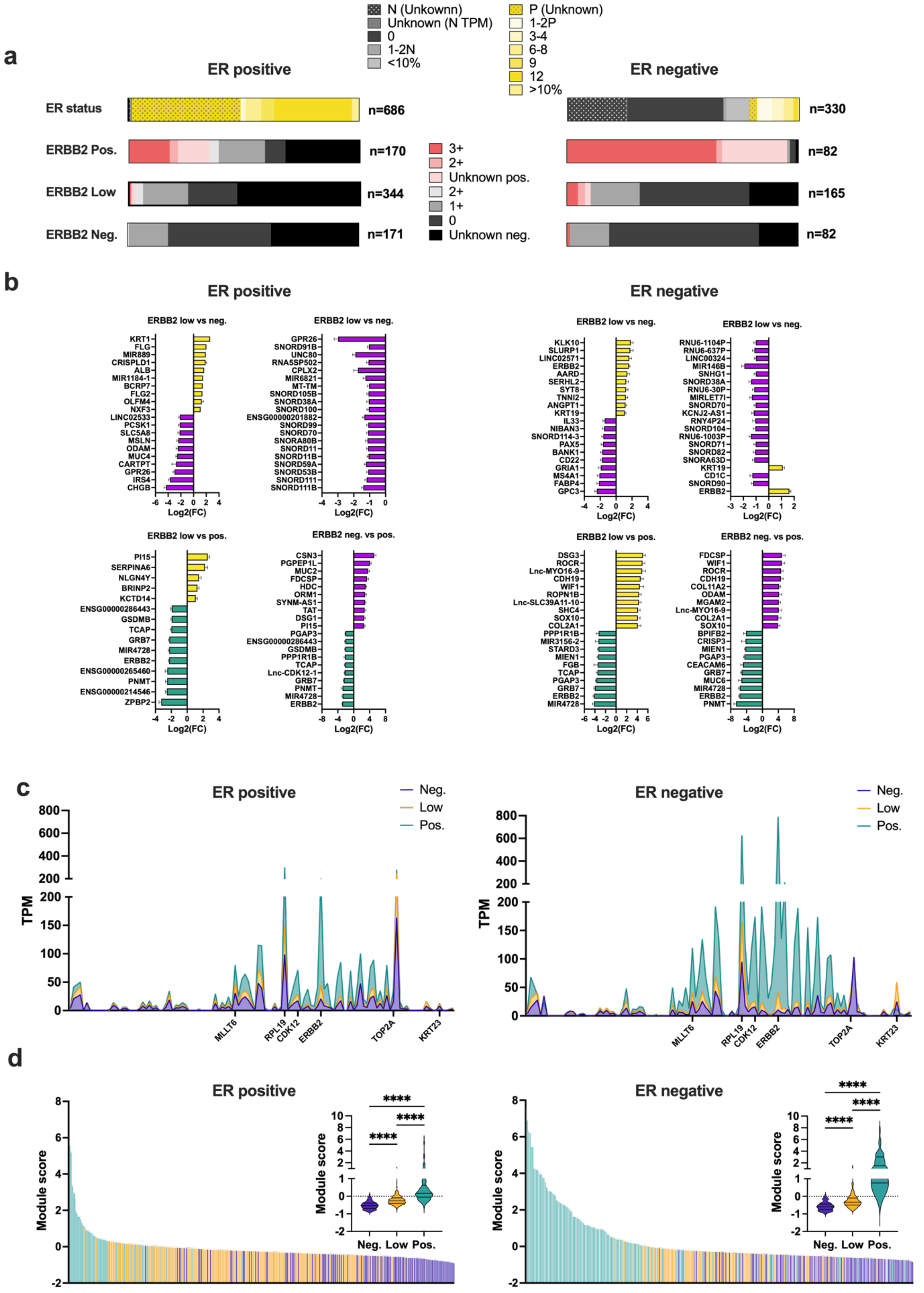

**Supplementary Figure 9. Differential gene expression analysis of *ERBB2* positive, low, and negative tumors.** (**a**) Overview of samples stratified into *ESR1* positive and negative groups in reference to the original IHC classifications from the biobanks. Each *ESR1* group was further divided into ERRB2 positive, low, and negative subgroups according to TPM interquartile ranges. Each bar shows the proportion of samples in each subgroup according to the original IHC classifications from the biobanks. **(b)** Bar graphs show differentially expressed genes between *ERBB2* expression groups. **(c)** Line plots compare gene expression between *ERBB2* expression groups across 115 genes located near the 17q11-21 locus (e.g,. extended Her2 amplicon). **(d)** Distribution of Her2 amplicon gene signature scores between *ERBB2* expression groups ranked from highest to lowest. Inset violin plots compare mean Her2 amplicon scores between *ERBB2* expression groups. Kruskal-Wallis test followed by Dunn's multiple comparisons test, ****P<0.0001.

**Supplementary Figure 10.**

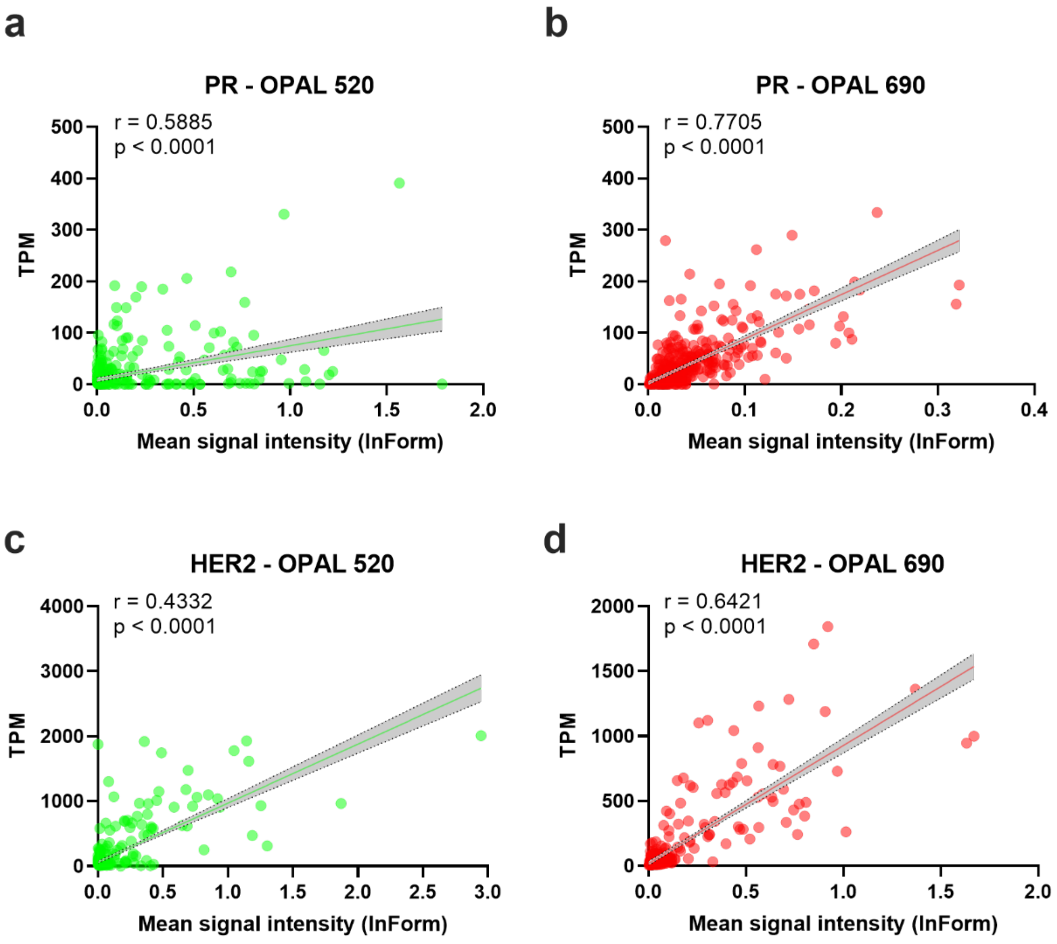


**Supplementary Figure 10. Correlations between RNA-FISH and RNA-SEQ improve when detecting biomarker signals at far red wavelengths.** Detection of (**a-b**) progesterone receptor (PR) and (**c-d**) HER2 with either OPAL 520 fluorophore (525 nm emission; **a**, **c**) or OPAL 690 fluorophore (694 nm emission; **b**, **d**). Spearman's rho correlation; 95% confidence interval of the best fit linear regression line shown in shaded region.

**Supplementary Table 1. Diagnostic performance of RNA-seq training and test data set**

| Marker | PR | | ER | | HER2 | | Ki-67 | |
|---|---|---|---|---|---|---|---|---|
| Train/Test data | Train | Test | Train | Test | Train | Test | Train | Test |
| TP | 393 | 165 | 486 | 212 | 89 | 40 | 106 | 45 |
| FP | 25 | 11 | 16 | 11 | 6 | 6 | 1 | 1 |
| TN | 238 | 104 | 171 | 71 | 578 | 258 | 49 | 14 |
| FN | 36 | 28 | 29 | 18 | 28 | 8 | 9 | 6 |
| Accuracy | 0.9118 | 0.8734 | 0.9359 | 0.9071 | 0.9515 | 0.9551 | 0.9394 | 0.8939 |
| 95% CI | 0.8882 | 0.831 | 0.9152 | 0.8692 | 0.9329 | 0.9259 | 0.8914 | 0.7936 |
| | 0.9319 | 0.9084 | 0.9529 | 0.9369 | 0.9662 | 0.9753 | 0.9706 | 0.9563 |
| No Information Rate | 0.6199 | 0.6266 | 0.7336 | 0.7372 | 0.8331 | 0.8462 | 0.697 | 0.7727 |
| P-Value [Acc >NIR] | <2e-16 | <2e-16 | <2e-16 | 4.35E-14 | <2.2e-16 | 9.42E-10 | 1.19E-14 | 9.51E-03 |
| Kappa | 0.8144 | 0.7372 | 0.8395 | 0.7665 | 0.8114 | 0.8247 | 0.8627 | 0.7298 |
| Mcnemar's Test P-Value | 0.2004 | 0.01041 | 0.07364 | 0.2652 | 0.0003164 | 0.7893 | 0.02686 | 0.13057 |
| Sensitivity | 0.9161 | 0.8549 | 0.9437 | 0.9217 | 0.7607 | 0.8333 | 0.9217 | 0.8824 |
| Specificity | 0.9049 | 0.9043 | 0.9144 | 0.8659 | 0.9897 | 0.9773 | 0.98 | 0.9333 |
| Pos Pred Value | 0.9402 | 0.9375 | 0.9681 | 0.9507 | 0.9368 | 0.8696 | 0.9907 | 0.9783 |
| Neg Pred Value | 0.8686 | 0.7879 | 0.855 | 0.7978 | 0.9538 | 0.9699 | 0.8448 | 0.7 |
| Precision | 0.9402 | 0.9375 | 0.9681 | 0.9507 | 0.9368 | 0.8696 | 0.9907 | 0.9783 |
| Recall | 0.9161 | 0.8549 | 0.9437 | 0.9217 | 0.7607 | 0.8333 | 0.9217 | 0.8824 |
| F1 | 0.928 | 0.8943 | 0.9558 | 0.936 | 0.8396 | 0.8511 | 0.955 | 0.9278 |
| Prevalence | 0.6199 | 0.6266 | 0.7336 | 0.7372 | 0.1669 | 0.1538 | 0.697 | 0.7727 |
| Detection Rate | 0.5679 | 0.5357 | 0.6923 | 0.6795 | 0.127 | 0.1282 | 0.6424 | 0.6818 |
| Detection Prevalence | 0.604 | 0.5714 | 0.7151 | 0.7147 | 0.1355 | 0.1474 | 0.6485 | 0.697 |
| Balanced Accuracy | 0.9105 | 0.8796 | 0.9291 | 0.8938 | 0.8752 | 0.9053 | 0.9509 | 0.9078 |

**Supplementary Methods.**

#### *Study Design*

Given the large number of samples, we divided the workflow into various batches of at least 8 samples (8-24 samples depending on the laboratory technique) that were nested into three layers of batches that followed a sequential order: 1) LCM (6-16 samples per batch, including samples with more than one sample dissected by LCM), 2) RNA isolation (8-24 samples), and 3) RNA-SEQ library preparation (8-46 samples). Batches earlier in the workflow were maintained in later batches. For example, two LCM batches could be combined into one RNA isolation batch and then two RNA isolation batches could be combined into one RNA-SEQ library preparation batch. No samples from a given batch were split into separate batches in the downstream workflow, unless a sample had to be redone due to technical issues.

#### *Tissue processing*

For tissues provided by PATH Biobank, sectioning was performed at MultiplexDX where several serial sections were obtained in the following order: 2 sections on Epredia Superfrost plus slides (one slide used for RNA-FISH and the second slide used as a backup for failed RNA-FISH runs), 1 section on a Leica PEN membrane slide (2 µm thick membrane and certified RNase-free) that was used for a rapid cresyl violet stain followed by laser capture microdissection, 1 section mounted on an Epredia Superfrost plus slide that was used for H&E staining and reviewed by a board certified pathologist, and 1 section was taken as a scroll, placed into a 1.5 mL microcentrifuge tube (Eppendorf DNA LoBind® Tubes, Cat. No. 0030108051), and frozen at –20 °C for later RNA extraction and RNA sequencing. To ensure enough sections in case of technical issues that led to failed assays or for follow up analyses, we took an additional second set of serial sections in the same order as described above. Sectioning was conducted in an RNase-free manner using a slightly modified protocol recommended by Leica 1 to ensure RNA integrity for laser capture microdissection and further molecular analysis of RNA species. Sections were stored in standard microscope slide boxes either at room temperature or the refrigerator until further analyses were performed. For tissues obtained from Biobank Graz, the tissue sectioning was performed at the Center for Medical Research at the Medical University of Graz, where 8 serial sections were obtained in the following order: 1st section on Epredia Superfrost plus slide, 2nd section on Epredia Superfrost plus slide, 3rd section on a Leica PEN membrane slide, 4th section on Epredia Superfrost plus slide, 5th section on Epredia Superfrost plus slide, 6th section on a Leica PEN membrane slide, 7th section on Epredia Superfrost plus, 8th section taken as a scroll/curl, placed into a 1.5 mL microcentrifuge tube (Eppendorf DNA LoBind® Tubes, Cat. No. 0030108051 or equivalent), and frozen at –20 °C for later RNA extraction and RNA-SEQ.

#### *Whole-slide imaging, image annotation, and image analysis*

The Vectra Polaris is the ideal setup for RNA-FISH images because the instrument contains proprietary filters that allow spectral unmixing of fluorophore signals as well as the ability to isolate unwanted autofluorescence. This alleviates two limitations of RNA-FISH: crosstalk/bleed-through fluorophore emission spectra (especially with high target expression) as well as background noise due to inherent autofluorescence (e.g., lipofuscin, collagen, blood cells) and autofluorescence from tissue preparation (e.g., formalin fixation).

During our selection process of the appropriate OPAL dyes for each target, we found that RNA-FISH biomarkers that were detected using Opal 520, a fluorescent dye that emits in the green wavelength (525 nm emission maxima), showed the poorest concordance with RNA-SEQ data that was independent of the marker (PR: r=0.59 and HER2: r=0.43; **Supplementary Figure 10a, c**). Switching the fluorophores of these markers to Opal 690 improved concordance substantially (PR: r=0.77 and HER2: r=0.64; **Supplementary Figure 10b, d**). This is likely due to the high levels of background and endogenous tissue autofluorescence that may arise from tissue processing. Because of this, we selected the highest expressed target *ERBB2* to be detected with Opal 520 to maximize the signal to noise.

To select regions of interest for LCM, we considered both the H&E-stained specimen and RNA-FISH biomarker expression to identify regions of interest (ROI) that displayed heterogeneity. This included specimens that contained more than one histological subtype (ductal carcinoma, lobular carcinoma, mucinous, etc.), the presence of DCIS/LCIS mixed with invasive BCa, or biomarker heterogeneity. Biomarker heterogeneity consisted of ROIs that were positive for a particular marker in one area, but negative for the same marker in another area on the same tissue section (i.e., all or none heterogeneity). Alternatively, we also considered heterogeneity in expression levels where a biomarker is highly expressed in one area but lowly expressed in another area on the same tissue section (i.e., high or low heterogeneity). In some instances, we also conducted LCM on both invasive BCa and adjacent healthy breast epithelial cells.

##### *RNA isolation and quality control*

The Macherey Nagel NucleoSpin totalRNA FFPE XS kit was used for RNA isolation using the manufacturer´s protocol with the following deviations as suggested by the user instructions: 1) omission of the deparaffinization step since this already occurs before cresyl violet staining; 2) lengthening proteinase K digestion to 90 min at 56 ºC; 3) inclusion of the optional on-column DNase digestion step to remove residual DNA. Laser capture microdissected samples were processed in batches of 8-16 samples with a single person conducting the isolation procedure for a single batch.

##### *RNA library preparation and sequencing*

We followed several of the manufacturer's recommended changes for using highly degraded FFPE specimens with the Takara SMARTer Stranded Total RNA-Seq Kit v3 - Pico Input Mammalian, including 1) total RNA input range from 0.5-15 ng, although we found the best performance to be from 1 to 20 ng with PCR cycles adjusted accordingly; 2) inclusion of the DNase pretreatment step 3) omission of the fragmentation step; 4) 10 PCR cycles for first PCR amplification step; and 5) 10-13 PCR cycles for the 2nd PCR amplification step depending on starting input. For 1 ng input, we used 13 PCR cycles and for 10 ng and 20 ng inputs, we used 10 PCR cycles. Libraries were prepared in batches of 8-48 samples by an individual scientist, while maintaining the batch organization of prior procedures like laser capture microdissection and RNA isolation.

The positive control sample consisted of a mixture of total RNA extracted from 3 biological replicates of each IHC-surrogate subtype (e.g., luminal A and B, HER2+, and TNBC), including both node-negative and node-positive specimens (24 specimens total). This cocktail positive control was aliquoted into single-use aliquots and one sample was prepared for each library preparation batch alongside all other samples. This positive control had a similar quality, quantity, and gene expression relative to our experimental samples and aided in identifying technical batch effects. A second control consisted of a panel of 92 non-overlapping synthetic spike-in RNA controls characterized by the External RNA Control Consortium (https://www.nist.gov/programs-projects/external-rna-controls-consortium) 2 and a mixture of 69 isoform transcripts (SIRV-Set 3; Lexogen, Vienna, Austria) that could be used to identify processing artifacts in individual samples and monitor the performance of library preparation (e.g., dynamic range, technical variability, the limit of detection, etc.). These synthetic spike-ins were introduced into every sample before library preparation at an attomolar input concentration that resulted in an estimated 1% of mRNA reads by using Lexogen's calculator worksheet.

The bioinformatics pipeline for RNA sequencing is outlined below. Details about statistical analyses for each pipeline can be found in their respective user manuals so it is only briefly summarized here:

1) FASTQC (v0.11.9) – used for initial quality check of raw sequencing data to provide general descriptive stats about sequence quality, identify duplicates, potential biases for downstream processing, sample outliers, and over-represented sequences/contaminants.

2) Cutadapt (v3.4) – Removal of adapters and filtering for sequence quality and length.

3) STAR (v2.7.7a) – alignment tool used to map reads to the human genome, providing metrics about mapped/unmapped reads, low-quality reads, and ambiguous reads.

4) SAMtools (v1.16.1) – used for indexing and sorting bam files.

5) UMI-tools (v1.0.0) – used for deduplicating bam files based on unique molecular identifiers introduced during library preparation.

6) FeatureCounts (v1.5.2) – this was used to identify gene and transcript abundance across various RNA biotypes (e.g., mRNA, snRNA, lincRNA, rRNA, etc.). Read counts were normalized to transcripts per million (TPM) for an overview of biotypes and transcript abundance.
7) DESeq2 (v.1.36.0) and Wilcoxon rank sum tests – Raw integer count matrices were imported into DeSeq2, a tool used to assess differential gene expression using a generalized linear model under consideration of a negative binomial distribution (Poisson-gamma model). To estimate differentially expressed genes, the shrinkage estimator apeglm was used. Wilcoxon rank sum tests were also used to test for differential expression in a non-parametric approach. To account for multiple hypothesis testing, FDR adjusted P-values ($P < 0.01$) were used to assess significance.

*Generation of molecular subtyping 293-gene list*

To generate a list of genes for molecular subtyping of samples into luminal A, luminal B, HER2, and basal-like subtypes, we conducted differential gene expression (DE) analysis using DESeq2 and the Wilcoxon rank-sum test [1], a nonparametric approach that better controls the false discovery rate (FDR) when analyzing large sample sizes. We assessed DE between each subtype combination (e.g., luminal A vs luminal B, luminal A vs Her2, etc.) and took the union of the top 50 significant DE genes (FDR adjusted $p < 0.05$ with fold change > 4), which yielded 196 genes (pseudogenes were manually removed). We generated a second list of genes to better differentiate high vs low OncotypeDX luminal samples using the following approach: First, we filtered luminal A and luminal B samples, which did not agree between the biobank surrogate subtypes based on St. Gallen criteria (e.g., Ki-67 < 30 for luminal A or Grade 3 for luminal B) and our classification based on 196 genes. Then we classified discordant luminal samples to either high or low risk based on their OncotypeDX score. Next, we performed DE analysis between luminal high and low risk samples by DESeq2 and Wilcoxon, took the intersected results from these two methods, and selected 30 significant DE genes (FDR adjusted $p < 0.05$ with fold change > 4; pseudogenes were manually removed). We also conducted unsupervised clustering and selected 78 genes that were enriched in the luminal androgen receptor (LAR) TNBC subtype. We then took the union of our 196 subtype DE genes, 30 luminal high/low risk genes, and 78 LAR TNBC subtype genes, to generate a final list of 293 genes. To classify individual samples, we used consensus clustering (kmeans = 4; ConsensusClusterPlus, v.1.60.0 package in R) with this 293-gene list. Heatmaps were generated by the ComplexHeatmap package in R (v.2.12.1).

*Benchmarking subtyping classifiers and development of consensus subtyping scheme*

The mFISHseq 293-gene molecular subtype classifier was benchmarked against the IHC-surrogate subtypes (using the definitions proposed by the 2013 St. Gallen International Breast Cancer Conference Panel [2]), research based PAM50 and AIMS implemented in the GeneFu package [3], as well as the six and four Lehmann's TNBC subtypes [4,5] using TNBCtype [6]. To assess risk of single samples belonging to a particular subtype, we again used GeneFu to derive scores for several research based prognostic signatures, including the 21-gene OncotypeDX, GENE70 (research version of Mammaprint), the PAM50 Risk of Recurrence by Subtype only (ROR-S), and Genomic Grade Index (GGI). For clinical risk, we used a modified Adjuvant! Online criteria outlined in the TAILORx [7] and MINDACT [8] trials to classify samples as low or high risk.

To derive a consensus subtype, we used a simple voting scheme between multigene subtyping tools (mFISHseq, PAM50, and AIMS) where the consensus subtype was determined by the majority. In seven cases (Luminal A: n=3; Luminal B: n=2; TNBC: n=2), the three multigene subtyping tools assigned a different subtype to each case, so we used the IHC-surrogate subtype as the tiebreaker. Other comparison metrics included concordance between all samples and stratified by subtype using Cohen's kappa statistics, survival using KM plots and log rank tests, and stability assessed by correlation to PAM50 centroids.

##### *Constructing consensus prognostic risk categories*

As described above, we used GeneFu to derive scores for several research based prognostic signatures (OncotypeDX, GENE70, ROR-S, and GGI) and clinical risk according to modified Adjuvant! Online criteria outlined in the TAILORx [7] and MINDACT [8] trials to classify samples as low, intermediate (OncotypeDX and ROR-S only) or high risk. For samples that did not have detailed enough information to use modified Adjuvant! Online criteria, we instead used the American Joint Committee on Cancer ($8^{th}$ edition, 2017) pathological prognostic stage groups with stage IA and IB classified as low risk and IIA and above classified as high risk. The high, low, and ultra-low consensus prognostic risk categories were defined according to **Supplementary Figure 8d.**

##### *Univariate and multivariate logistic regression using I-SPY2 trial data*

We downloaded data from the T–DM1, control, and pertuzumab arms of the I-SPY2 trial from NCBI's Gene Expression Omnibus (GEO) under accession code GSE181574. The T-DM1 arm contains 52 patients (ER+/HER2+: n=35, ER–/HER2+: n=17) treated with T-DM1 and pertuzumab with 30 patients achieving pathological complete response (pCR). The control arm contains 31 patients (ER+/HER2+: n=19, ER–/HER2+: n=12) treated with paclitaxel and trastuzumab with 8 patients achieving pCR. The pertuzumab arm (used as a control arm for pertuzumab in the T-DM1 arm) contains 44 patients (ER+/HER2+: n=29, ER–/HER2+: n=15) treated with paclitaxel, pertuzumab, and trastuzumab with 26 patients achieving pCR. The T-DM1 dataset was stratified into training and test (50:50) data based on pCR and hormonal status. To associate the gene/gene signature with pCR, we performed univariate logistic regression in R through lmtest (v09-40) with the likelihood ratio test assessing significance. We further combined genes and gene signatures into the multivariate logistic model with elastic net regularization using tidymodels (v 1.1.1) with hyperparameter tuning by 10-fold cross validation. The final 19-feature classifier (called multivariate T-DM1_pred) was then applied to the T-DM1 test set and compared with ERBB2 alone. We also constructed a single score (called univariate T-DM1_pred) by taking the individual features and multiplying them by a weighted coefficient, which was determined by multiplying the beta coefficient from the univariate analysis by the log of the p-value (e.g., univariate T-DM1_pred score = sign((beta coefficient)*log(p))*feature 1 expression value + sign((beta coefficient)*log(p))*feature 2 expression value + …). The sum of the weighted value of each feature was then averaged and transformed into z-scores.
